# Comparison of long-read sequencing strategies for resolving complex genotypes at Facioscapulohumeral dystrophy-associated loci

**DOI:** 10.1101/2025.08.08.25333030

**Authors:** Charlotte Tardy, Jean Philippe Trani, Victor Murcia Pienkowski, Loeva Morin, Christel Castro, Louis Souville, Camille Humbert, Laurène Gérard, Nathalie Eudes, Amire Assoumani, Karine Bertaux, Camille Verebi, Juliette Nectoux, Emmanuelle Salort Campana, Marie Line Jacquemont, Annick Toutain, Martial Mallaret, Céline Tard, Mélanie Fradin, Shahram Attarian, Karine Nguyen, Rafaëlle Bernard, Frédérique Magdinier

## Abstract

**Background:** Facioscapulohumeral dystrophy (FSHD) is typically caused by contraction of the D4Z4 repeat array at chromosome 4q35 (FSHD1) or pathogenic variants in the SMCHD1 gene (FSHD2). While these account for the majority of cases, 1–2% of patients present with clinical features of FSHD but lack a known genetic cause, revealing a diagnostic gap. In Previous studies, we identified over 70 patients with structural rearrangements involving the 4q35 or 10q26 loci, some of which may be pathogenic in the absence of other FSHD-associated features.

**Methods:** Given their diagnostic relevance, we performed detailed structural analyses of these rearrangements using high-resolution long-read sequencing technologies (Oxford Nanopore and PacBio) for seven patients carrying different structural variants of the 4q35 or 10q26 loci.

**Results:** These approaches enabled us to resolve nucleotide-level architecture and methylation patterns across the 4q35 and 10q26 loci. We show that duplicated alleles arise from intrachromosomal recombination between LSau and β-satellite elements, producing variable deletions within D4Z4, with breakpoints differing among patients. These complex structural variants are not detectable using standard technologies like Bionano Optical Genome Mapping and require manual curation for identification. Importantly, determining the pathogenic relevance of these rearrangements necessitates integrating structural and epigenetic features typically associated with FSHD.

**Conclusion:** The results highlight the importance of thorough molecular analysis for FSHD patients who test negative for FSHD1 and FSHD2, advocating for expanded diagnostic strategies. Comprehensive evaluation of 4q35 structural variants is essential for improving diagnosis accuracy, guiding genetic counseling, and optimizing care for patients with atypical FSHD presentations.

**Graphical Abstract:** Hypothetical model for the formation of 4q35 intrachromosomal duplications. We speculate that the breakpoint occurs in the proximal region of a D4Z4 units that corresponds to LSau repeats and involves the distal βsatellite array. The duplicated array is then inserted in the distal part of the locus mapped by the most telomeric red probe used for MC.

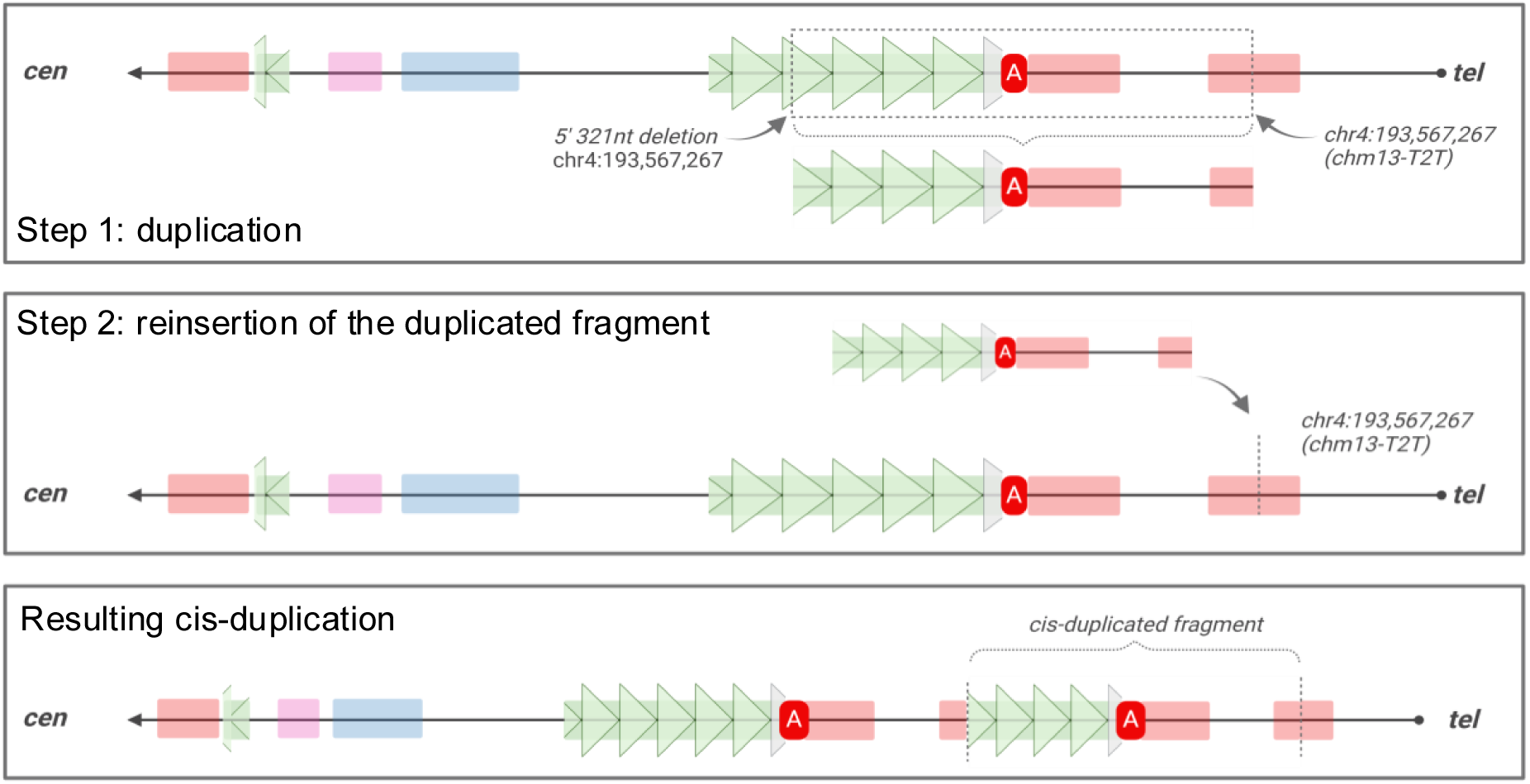

## Background

With a prevalence between 1/8,000 and 1/12,000 (1–3), Facioscapulohumeral Dystrophy (FSHD) is the third most common muscular dystrophy. It is clinically defined by a progressive and asymmetric muscle weakening usually starting in the second decade of life that affects specific muscles of the face and scapular girdle. During late-stage progression, the disease may gradually affect the pelvic girdle and lower limb muscles, resulting in a significant motor disability (2). About 20% of patients with a classical FSHD phenotype become wheelchair users at late progression stages. FSHD exhibits a wide clinical variability among individuals, with variable penetrance and expressivity, including within families with a shared molecular alteration (4).

FSHD is linked to the subtelomeric 4q35 locus (5, 6). It is estimated that 95% of clinical cases (FSHD1; MIM#158900) present a contraction of the D4Z4 macrosatellite array with patients carrying between 1 to 10 units of the repeat at the 4q35 locus. In the control population wild-type alleles usually range from 11 to up to 150 D4Z4 units (6–8).

Each D4Z4 repeat is 3.3 kb in size and contains >70% of GCs with >65% of them being methylated in the healthy population but hypomethylated in FSHD patients (9, 10). The pathological contracted allele is also associated at its 3’ end with a type A haplotype (dubbed pLAM) that contains a ATTAAA polyadenylation signal (PAS) required for stabilization of the *DUX4* mRNA transcribed from the last D4Z4 unit (11, 12). In addition, microsatellites and Simple Sequence Length Polymorphisms (SSLP) of different sizes are present proximal to D4Z4 with several of these short tandem repeats being more frequent in FSHD patients (13). In most cases, FSHD is inherited as an autosomal dominant trait. Between 10-30% of cases are sporadic, including instances of somatic mosaicism, which account for 40% of the *de novo* FSHD cases (14, 15). Mosaicism was reported in 14% of non-affected parents, but also in 3% of the general population (16, 17). In FSHD1, the severity of the disease roughly and inversely correlates with the number of D4Z4 units in FSHD1, although several exceptions exist (4).

Due to duplication events during evolution, the subtelomeric 4q and 10q regions are 98% homologous and interchromosomal rearrangements between these two regions lead to atypical 4q/10q hybrid D4Z4 arrays in at least 10% of the European or Asian populations (17–19). However, 10q alleles are not associated with the pathology (20–22), and only short alleles on 4q coexist with the disease regardless of the origin of the D4Z4 repeats, *i.e* 4q- or 10q-type (18, 22). This implies a positional effect of D4Z4 contraction on chromosome 4q rather than an importance on the type of D4Z4.

Type 2 FSHD (FSHD2; MIM#158901, 5% of patients) is phenotypically identical to FSHD1 but has a digenic mechanism involving the presence of a permissive A-type haplotype on a > 11 D4Z4 alleles and the presence of a variant in the *SMCHD1* gene encoding a chromatin-associated factor (23). Loss of function or haploinsufficiency in SMCHD1 result in a marked hypomethylation of D4Z4 (9, 23). Recently, pathogenic variants of *DNMT3B* and *LRIF1* genes were associated with rare cases of FSHD (24, 25) but in 1-2% of patients presenting typical clinical features of FSHD, the cause of the disease remains unknown.

In most laboratories worldwide, the diagnosis of FSHD is performed by Southern Blotting (SB) after digestion of genomic blood DNA with the *Eco*RI enzyme, linear or pulsed-field electrophoresis, and hybridization with the p13E-11 probe that maps the D4F104S1 marker located upstream of the first D4Z4 unit (7, 26). Restriction enzymes specific to D4Z4 repeats on 4q (*Xap*I/*Apo*I) or 10q (*Bln*I/*Avr*II) might also be used in combination with *Eco*RI to distinguish between the two types of alleles (27). However, in approximately 20% of cases, molecular diagnosis by SB is hindered by technical difficulties inherent to the methodology or the quality of the DNA, as well as by 4q-10q translocations, somatic mosaicism, or more complex rearrangements. More recently, novel technologies were developed such as Molecular Combing (MC) (28) or Single Molecule Optical Genome Mapping (OGM) (29, 30). MC, based on *in situ* hybridization allowed, a complete analysis of each 4q and 10q allele, the sizing of the number of D4Z4 repeats and the identification of the associated A or B type distal haplotypes in a single step (28). OGM, recently developed for visualization of structural variation of the genome (SVs), is based on imaging of long labeled DNA fragments (29, 30). As alternative and promising emerging methods, Long Read sequencing techniques, provided by Oxford Nanopore Technology (ONT) or PacBio Hifi sequencing now permit sequencing of long DNA fragments together with profiling DNA methylation. ONT operates by detecting signal variations as a single-stranded DNA passes through a biological nanopore, enabling the production of very long reads that can exceed 1 Mb (31). It is particularly useful and appropriate for analyzing structural variants such as long deletions in FSHD (32) where short read sequencing typically generates reads that do not exceed 500 bases. Reads generated by HiFi PacBio circular consensus sequencing (CCS) technology are fragmented to be only around 30 kb in size, but with a higher sequencing accuracy compared to ONT and a mean error rate of 0.0007% compared to 0.0042% for ONT most recent R10.3 flow cell (33).

Using MC, we previously reported the existence of complex chromosomal rearrangements at the 4q35 and 10q26 loci in patients with clinical signs evocative of FSHD (34–36). The most common of these rearrangements that we identified in more than 30 patients(34, 35), consist in a repetitive pattern of two D4Z4 arrays of different sizes, duplicated *in cis* and carried by the same chromosome. These *cis-*duplicated arrays contain a variable number of D4Z4 units and are flanked distally by a type A haplotype (34). In most reported cases, the proximal array is large (>11 D4Z4 units), while the distal array is contracted (less than 25 kb; 11 units). The association of these alleles with FSHD is not clearly established, as they do not always segregate with clinical signs of the disease within families (19, 27, 36). It was suggested that *cis*-duplicated alleles could be involved in a digenic transmission, as they are often associated with pathogenic *SMCHD1* variants (35) or the presence of a short D4Z4 allele on the other 4q chromosome (35). However, in approximately 1/3 of cases, the *cis*-duplicated allele is the only genetic alteration suggesting that it might also be causative of the disease (34, 35, 37). Large deletions encompassing part of the D4Z4 array and the proximal region upstream of D4Z4 that hybridizes the p13E-11 probe, as well as complex rearrangements of 10q alleles have also been documented in individual clinically diagnosed with FSHD (34, 36). Further adding on the complexity in characterizing these complex SVs, the assembly of homologous 4q and 10q loci and respective haplotypes remain partial in the most recent T2T human genome releases (38).

From a medical perspective, it is essential to accurately characterize these rearrangements, not only to understand their role in FSHD pathogenesis, but more importantly, to provide answers to patients who currently remain undiagnosed.

To this aim, we leveraged the high resolution offered by various long-read sequencing technologies to investigate the nucleotide-level organization of the 4q35 locus in FSHD patients carrying complex genomic rearrangements. Specifically, we performed a comprehensive analysis of the 4q and 10q subtelomeric regions by comparing the performance Oxford Nanopore nCATS, Adaptive Sampling, and Whole Genome Sequencing (WGS), in parallel with PacBio WGS and Optical Genome Mapping (OGM, Bionano). This analysis was conducted on seven patients previously characterized by MC as carrying rearrangements at the 4q35 or 10q26 locus (34, 35). Our results reveal that duplicated alleles originate from intrachromosomal rearrangements involving LSau and β-satellite elements, leading to deletions of variable length and distinct breakpoint locations across patients. Ultimately, determining the pathogenic relevance of these rearrangements required further evaluation of additional disease-associated features.

## Results

### Comprehensive analysis of distal 4q and 10q subtelomeres for a custom-made reference assembly

Due to their inherent structural complexity, the hg37 and hg38 genome assemblies reported only partial sequences of 4q and 10q haplotypes. More recently, development of long fragment sequencing techniques has enabled the resolution of numerous gaps in the sequencing of the human genome (38). After filling the gaps of GRCh38 using ultra-long (>100kb) ONT reads to facilitate assembly and 20kb circular HiFi PacBio reads to reduce sequencing error rates, the Telomere-to-Telomere (T2T) Consortium published the first complete human genome sequence in 2022 (38). This current reference genome (CHM13-T2T v2.0) that derives from a complete hydatidiform mole carrying a haploid set of chromosomes with 4qA and 10qA haplotypes was the first assembly to unveil the sequence distal to the last D4Z4 repeat toward the telomeric end at 4q and 10q loci. In addition, the T2T consortium also released the sequencing of the diploid HG002 genome carrying a maternal 4qA and 10qA alleles, and a paternal 4qB and 10qA haplotypes (39). Up to now 10qB alleles that are rare in the population remain absent from genome releases.

Using the CHM13-T2T reference genome which includes 33 complete D4Z4 repeats on chromosome 4 (chr4:193,434,245–193,543,060) and 33 complete D4Z4 repeats on chromosome 10 (chr10:134,616,734–134,725,983), we first mapped the position of all MC probes for 4q and 10q loci (**Table S1, Figure 1A, Figure S1A**) to subsequently guide our comprehensive analyses of long read sequencing compared to MC images for patients with complex rearrangements. We determined the position of four arbitrarily designed regions to build a custom reference genome for each patient based on MC barcode. Region 1 maps 128kb upstream of the first D4Z4 unit (chr4:193,305,855-193,434,244) (**Figure S1B**). Region 2 corresponds to one D4Z4 unit that we duplicated according to the number of repeated units determined by MC (chr4:193,434,245-193,437,541) (**Figure S1C**). Region 3 encompasses the sequence downstream of the last D4Z4, the pLAM element and downstream type A haplotype (chr4:193,543,061-193,567,267) (**Figure S1D**). Region 4 (chr4:193,567,268-193,574,945) encompasses the most distal red probe used for MC and the telomeric end of the 4q35 locus. For sequencing analyses, the combined sequences were formatted and converted into an output FASTA file.

**Figure 1.**
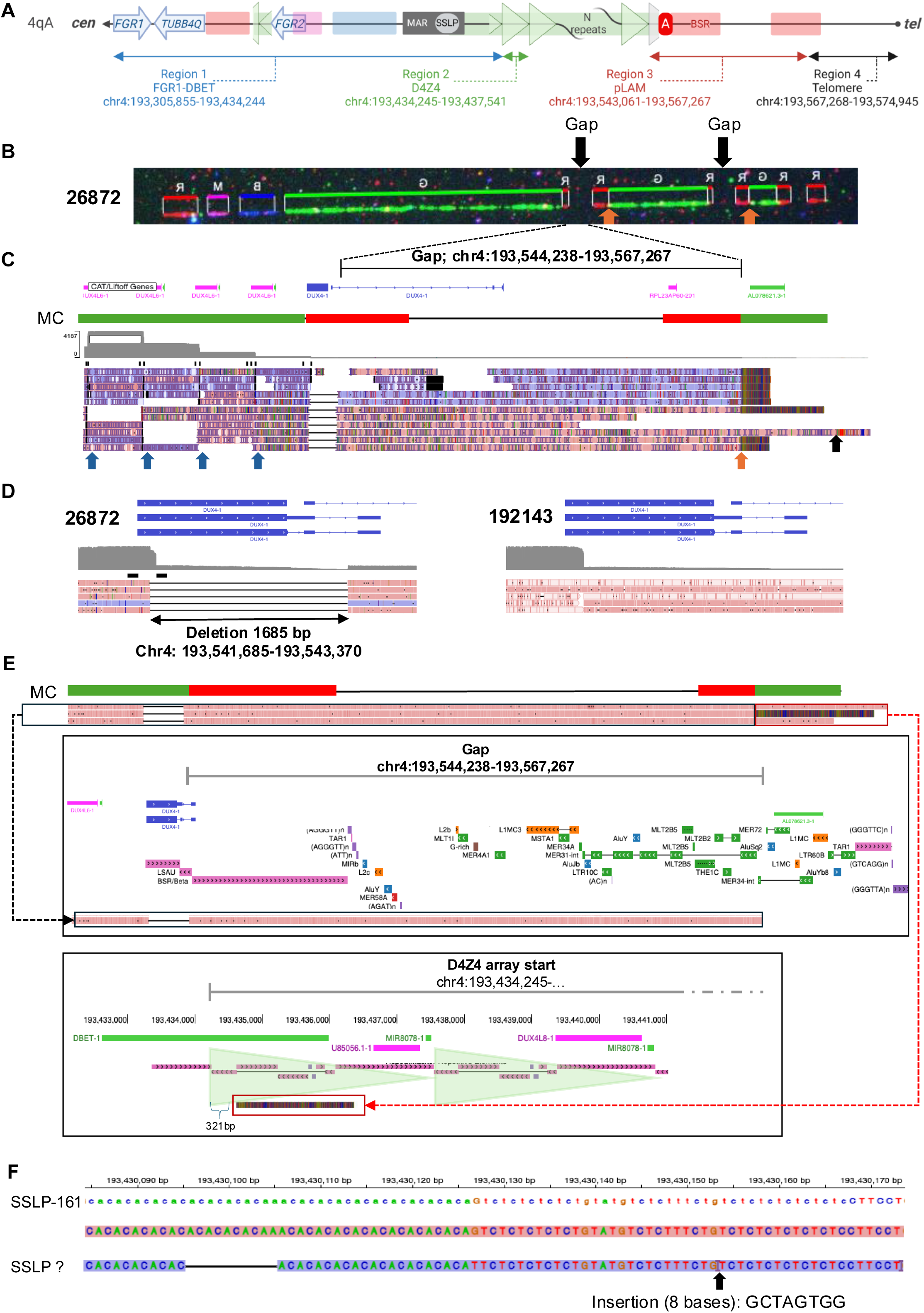
Characterization of *cis*-triplicated D4Z4 arrays by ONT. **A.** Schematic representation of the 4q35 region with position of different regions of interest relative to the CMH13-T2T genome assembly. The color code corresponds to the different probes used for molecular combing (MC). **B**. Raw image from MC for patient 26872. From left to right: the proximal red probe is specific to chromosome 4. The pink probe maps the proximal region that is shared between chromosomes 4 and 10. The blue probe maps to the p13E11 region. The green probe corresponds to D4Z4. Downstream of the green probe, the red probe corresponds to the qA haplotype. The first D4Z4 array (39 repeated units, RUs) is followed by two additional D4Z4 arrays (14 RUs and 5 RUs), each flanked by red probes. The most distal red probe on the right corresponds to the distal part of the 4q35 subtelomeric region. The “gap” is indicated by a black arrow. The fusion of signal between the D4Z4 and the subtelomeric probes is indicated by an orange arrow **C**. Visualization using IGV (Integrated Genome Viewer) of Oxford nanopore sequencing data. From top to bottom: RefSeq track, corresponding MC bar code, read coverage (ranging from 0 to 4187 reads), and aligned sequences. Blue arrows indicate the positions where CRISPR cuts were made using a D4Z4 gRNA. The orange arrow marks the breakpoint located at chr4:193,567,267, followed by a split read. The black arrow shows a read that starts at the last D4Z4 repeat and extends to the telomeric end. **D**. Close-up view of the DUX4 sequence. For patient 26872, the 4qA-L haplotype that corresponds to the presence of a longer final intron, is visible as a 1685 bp deletion when reads are compared to both the T2T reference genome and control sample 192143, who carries 4qA-S haplotypes. **E**. Schematic representation of the gap region (i.e. region that is not covered by MC probes; Chr4: 193,544,238-193,567,267) at the 4q35 locus. From top to bottom: representation of MC probes at this locus, one split-read showing the breakpoint at Chr4: 193,567,267, RefSeq Track and RepeatMasker Track from UCSC annotating repeated sequences. The black box highlights the portion of the gap region that aligns to the T2T reference sequence up to position 193,567,267. The red box indicates the poorly aligned region (split read). This sequence aligns to the beginning of a truncated 321 bp D4Z4 repeat unit. **F**. IGV view of the proximal polymorphic SSLP region spanning chr4:193,430,085-193,430,173 relative to the T2T CHM13 reference sequence (upper panel). Middle, patient 26872 carrie a SSLP-161 (CA10 AA CA10 GT CT5 GT AT GT CT2 TT CT GT CT6) allele and an atypical allele with homologies to the 4B162 haplotype (CA5 CA CA10 TT CT5 GT AT GT CT2 TT CT G GCTAGTGG T CT6) that contains an insertion of 8 bases (GCTAGTGG) (lower panel).

### Resolving D4Z4 duplicated arrays locus by ONT long-read sequencing and mapping to the T2T genome assembly

With the goal of providing a complete and comprehensive analysis of complex 4q35 rearrangements in patients affected with FSHD, we then evaluated the performance of different strategies for ONT long read sequencing of the distal 4q recombined alleles by selecting seven patients carrying different rearrangements (32, 34, 36) and a control individual (192143) (**Figure S2A**, **Table S2**).

The first patient (26872), is in his 10’s and was referred with a history of muscle weakness that began in his first decade of life. He presents with an atypical muscular phenotype, that is not specific to FSHD. He exhibits weakness in facial muscles as well as involvement of the lower limbs. MC identified a 4qB allele with 36 repeated units (RUs) and a rearranged 4qA allele consisting of three consecutive arrays of 39, 14, and 5 D4Z4 RUs. The different arrays are separated by a so-called “gap” corresponding to a region that is not covered by MC probes (**Figure 1B**, black arrow). The duplicated arrays are flanked by red probes (**Figure 1B**). This patient also carries a short 10qA allele with 6 RUs and a 10qB allele with 24 RUs (34).

To more specifically target the rearranged locus and the “gap” region, we first applied a Cas9 targeted approach using a guide RNA targeting D4Z4 to increase the number of reads covering this “gaps”. Using this guide, 3.3 kb D4Z4-cut reads were preferentially sequenced by nanopores but we also obtained a significant number of off-target reads due to the presence of divergent D4Z4 sequences in the genome (**Figure S2B**), with 42% of reads being on-target.

Given the length of the sequenced fragments (N50 value of 11.47 kb; maximum read length of 112 kb, **Sup Table S1**), it was challenging to conclusively differentiate the three D4Z4 arrays. Moreover, using the D4Z4 guide, we were not able to fully cover the last shortest triplicated array (5 RUs). However, we succeeded in sequencing the gap region entirely (**Figure 1C**). As observed by MC, this patient carries a A-type haplotype that we further characterized as an “L” subtype. Only the 4qS variant which is the main allele found in the European population is present in the current CHM13-T2T assembly (**Figure S3A**). By ONT, the 4qA-L haplotype is visible as a 1685 bp deletion spanning chr4: 193,541,685-193,543,370 that corresponds to intron 2, distal to the last D4Z4 unit (**Figure 1D, Figure S3A**) compared to the control (192143) who carries a 4qA-S haplotype (**Figure 1D**).

The three rearranged arrays are 100% homologous and all associated with a distal 4qA-L haplotype. Each duplicated D4Z4 array contains the restriction site for the *Xap*I enzyme, that is specific to chromosome 4 (**Sup Table S7**). We concluded that the triplication is caused by an intrachromosomal rearrangement, as the other 4q allele carried by this patient is a 4qB haplotype. In the most distal duplicated arrays (14 RUs; 5 RUs), the rearrangement is associated with the deletion of 321 bp of the proximal end of the first D4Z4 unit (position chr4:193,434,245-193,434,557) (**Figure 1E**). This identified breakpoint is consistent with that described by Lemmers *et al* (40) in two patients carrying a *cis*-duplication and incriminates LSau/β satellite elements in these recombination events through a common mechanism that occurs in *cis.* The breakpoint is located in a MER-34-int element, 280bp in 5’ of an AluSq2 element (**Figure 1E**). Using a reporter assay, we observed the absence of promoter or enhancer activity of this element when placed next to a reporter gene, compared to the DR1 sequence or empty vector (**Figure S3D**).

Mapping and sequencing of the most distal region of the locus, from the pLAM up to the first T_2_AG_3_ telomeric repeats showed a 98.4% homology when compared to the CHM13-T2T version 2.0 assembly (38). The so-called “gaps”, *i.e*. regions that are not covered by MC probes and located between duplicated arrays (**Figure 1B**, black arrows) span 23 kb in length and are homologous to the distal 4qA region (chr4:193,544,238-193,567,267). The “gap” region contains an intricate chain of mobile repeated elements such as Line, Alu, and endogenous retroviral sequences that are known to facilitate intra- or inter-chromosomal rearrangements (**Figure 1E, Figure S4A, B**). This result further confirm a *cis* rearrangements, without any inversion, contrary to our previous hypothesis (34).

In this patient, we also studied the proximal SSLP microsatellite (chr4:193,430,085-193,430,167 CHM13-T2T). He carries a SSLP-161 and an SSLP similar to 4B162 (41). However, this result requires confirmation due to the nanopore sequencing quality (**Figure 1F**). We also captured the complete short 10qA allele (6RU), characterized by an A-type haplotype containing the ATCAAA sequence instead of the 4qA-specific pLAM sequence (ATTAAA). The absence of a polyadenylation site suggests that the 10qA allele is non-pathogenic in this patient. Additionally, the presence of 6 *Bln*I restriction sites indicates the absence of 4q–10q rearrangement.

However, Despite the use of a patient-specific reference, the alignment quality remains poor due to the repetitive nature and limited length of the Nanopore reads, resulting in multiple ambiguous mappings within each D4Z4 array and low mapping quality scores (reads shown in white, **Figure S3B**). Due to the important dispersion of D4Z4 throughout the whole genome and subsequent misalignement, leading to a poor mapping, the lack of coverage over the most distal repeat array further illustrates the limitations of this strategy and ultimately led us to discard this approach based on the use of a single gRNA for precise allele characterization.

### Combination of guides optimize the sequencing strategy for identification of *cis*-rearrangements

To further improve the sequencing of patients carrying 4q35 rearrangements, we next combined our D4Z4 guide together with previously described p13E11 and pLAM guides (40, 42, 43) (**Sup Table S3**; **Figure S3C**). The second patient to be analyzed, is a female diagnosed during infancy. She shows a classical FSHD phenotype and has been explored by short read whole genome sequencing that came back negative for any differential diagnosis. By MC, we identified a *cis*-duplicated 4qA allele, with a long fragment consisting of 55 D4Z4 RUs, followed by a short fragment containing 1 D4Z4 unit (27858, **Figure 2A**). She also carries a 4qB allele of 13 RUs, a 10qA allele with 7 RUs, and another 10qA allele with 25 RUs (34).

**Figure 2.**
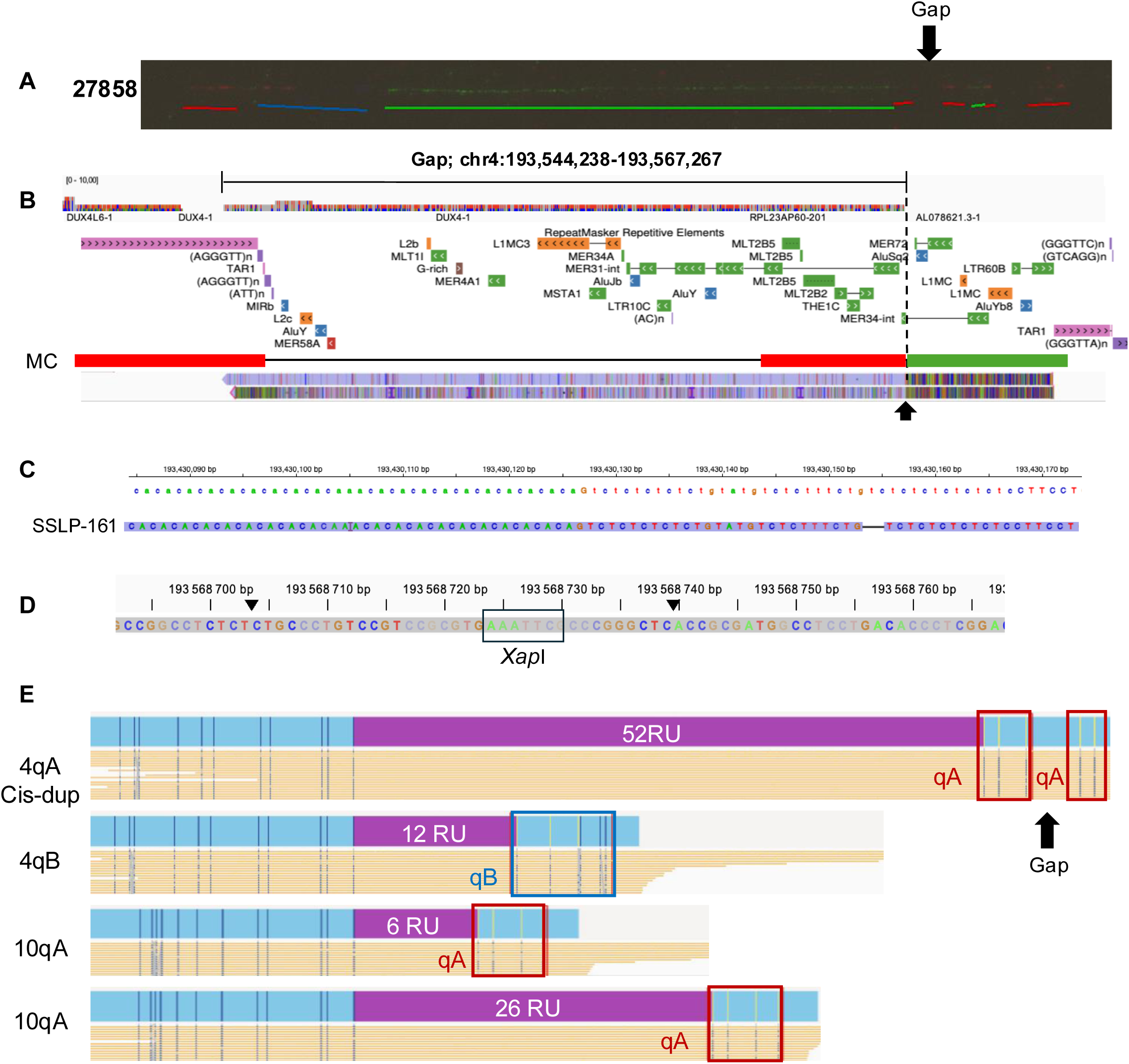
Comparison between MC, ONT and OGM for precise sizing of *cis*-duplicated D4Z4 arrays. **A.** Raw MC image for patient 27858. From left to right: the proximal red probe is specific to chromosome 4. The pink probe maps the proximal region that is shared between chromosomes 4 and 10. The blue probe maps to the p13E11 region. The green probe corresponds to D4Z4. Downstream of the green probe, the red probe corresponds to the qA haplotype. The first D4Z4 array (55 repeated units, RUs) is followed by a 1-unit fragment. The most distal red probe corresponds to the distal part of the 4q35 subtelomere. **B.** From top to bottom, sequencing coverage ranges from 0 to 10 reads. Representation of Repeat Masker track from UCSC with localization of MC probe, Lower panel, IGV snapshot showing the gap region (Chr4: 193,544,238-193,567,267) with the breakpoint corresponding to the *cis*-duplication at chr4:193,567,267 indicated by a black arrow. **C.** IGV view of the proximal polymorphic SSLP region spanning chr4:193,430,085-193,430,173 relative to the T2T CHM13 reference sequence. The patient carries a SSLP-161 (CA10 AA CA10 GT CT5 GT AT GT CT2 TT CT GT CT6) allele. **D.** IGV Snapshot of the *cis*-duplicated read showing a AAATTC *Xap*l restriction site located a position +1457 of the D4Z4 array (Black box). **E.** Bionano Optical Genome mapping (OGM) followed by FSHD EnFocus analysis of the 4q35 region for patient 27858. The D4Z4 array is represented in purple. The *cis*-duplicated allele was estimated to be 52 repeat units (RU) in size. A *cis*-duplication was suspected in the presence of an atypical incomplete A-haplotype pattern downstream of the D4Z4 array (red box). The gap region is represented in a black arrow. Using manual curation, the estimated size between the two unusual barcode was 16.2 kb (corresponding to 1 RU seen by MC). This patient, also carries a 4qB allele estimated of 12 RU (blue box), a 10qA at 6 RU and one at 26 RU, consistent with MC data.

A combination of three guides for ONT sequencing (**Figure S3C**) decreased the rate of on-target reads (16%) compared to the first sample (**Table S1)** and also increased the N50 value to 20 kb and the maximum read length to 164 kb. However, we obtained a low coverage and reduced yield due a lower quantity of long fragment DNA obtained after extraction from the plugs used for MC.

As for the previous patient, we also identified the presence of a 4qA-L haplotype on chromosome 4 together with the deletion of 321 bp corresponding to the LSau/β satellite elements located in the proximal part of each macrosatellite in this second patient. The distal breakpoint at chr4:193,567,267 is identical in the two patients who carry either a *cis*-duplication or -triplication (26872) and maps to the MER-34-int element, 280bp in 5’ of AluSq2 elements (**Figure 2B**). However, this breakpoint was initially missed by variant callers such as SVIM, NanoSV, and Sniffles2 (**Tables S4, S5 and S6**), probably because of the low depth of coverage at this locus with only 24 reads.

Mapping and sequencing of the gap region, from pLAM up to the breakpoint showed a 97.4% homology when compared to the CHM13-T2T version 2.0 assembly. This patient also carries a homozygous SSLP-161 haplotype as determined by ONT (**Figure 2C**). We also noticed the absence of *Bln*I and the presence of *Xapl* restriction sites in the duplicated repeat further supporting an intra-chromosomal 4q rearrangement (**Figure 2D, Table S7**).

For this patient, Bionano OGM was conducted in parallel and data were analyzed through the FSHD EnFocus Pipeline. The *cis*-duplicated array was not called by the FSHD EnFocus analysis tool but was detected by visual report curation as an atypical barcode, with an incomplete signal of the qA haplotype present in two copies, one following the other (**Figure 2E**). The estimated size between the two unusual barcode was 16.2 kb. OGM cannot give any information about the nature of distal duplicated region nor estimate the number of potential units. The size of the distal *cis*-duplicated array was estimated to one unit as found by MC and further confirmed by ONT sequencing (**Figure 2A**). As previously reported for other patients (34), this observation confirms a lower level of resolution of the OGM technique in cases of complex rearrangements and the need of manual data analysis in the presence of an atypical pattern.

### Overcoming Crispr-induced fragmentation by using an adaptive sampling strategy

The CRISPR-based approach causes breaks in the D4Z4 region due to the use of guides to enrich in sequences of interest, thus preventing a continuous sequencing of the locus in patients with large rearranged regions. To overcome this limitation, we next applied ONT Adaptive Sampling as an alternative ONT sequencing strategy. Adaptive Sampling selectively enriches or depletes specific DNA regions during sequencing by dynamically adjusting which fragments are retained or discarded from the sequencing pore, based on positions relative to the genome of reference.

Using this methodology, we first reanalyzed patient 26872 who carries a *cis*-triplicated 4qA allele (**Figure 3A**). For this patient, sequences were analyzed using the T2T diploid HG002 assembly to integrate the qA (maternal) and qB (paternal) haplotypes in the analysis. A total of 6 µg of DNA was needed for library preparation. Compared to adaptive sampling, the CRISPR/Cas method achieved a higher on-target read percentage (42% vs. 9%) and longer average read lengths (N50 of 11.47 kb vs. 452 bp, respectively). We confirmed what was observed with the Crispr-Cas approach, *i.e*, the presence of a 4qA-L homozygous haplotype, a “gap” region followed by a distal breakpoint at chr4: 193,567,267 and the presence of the 161 SSLP microsatellite array (**Figure 3B,C**).

**Figure 3.**
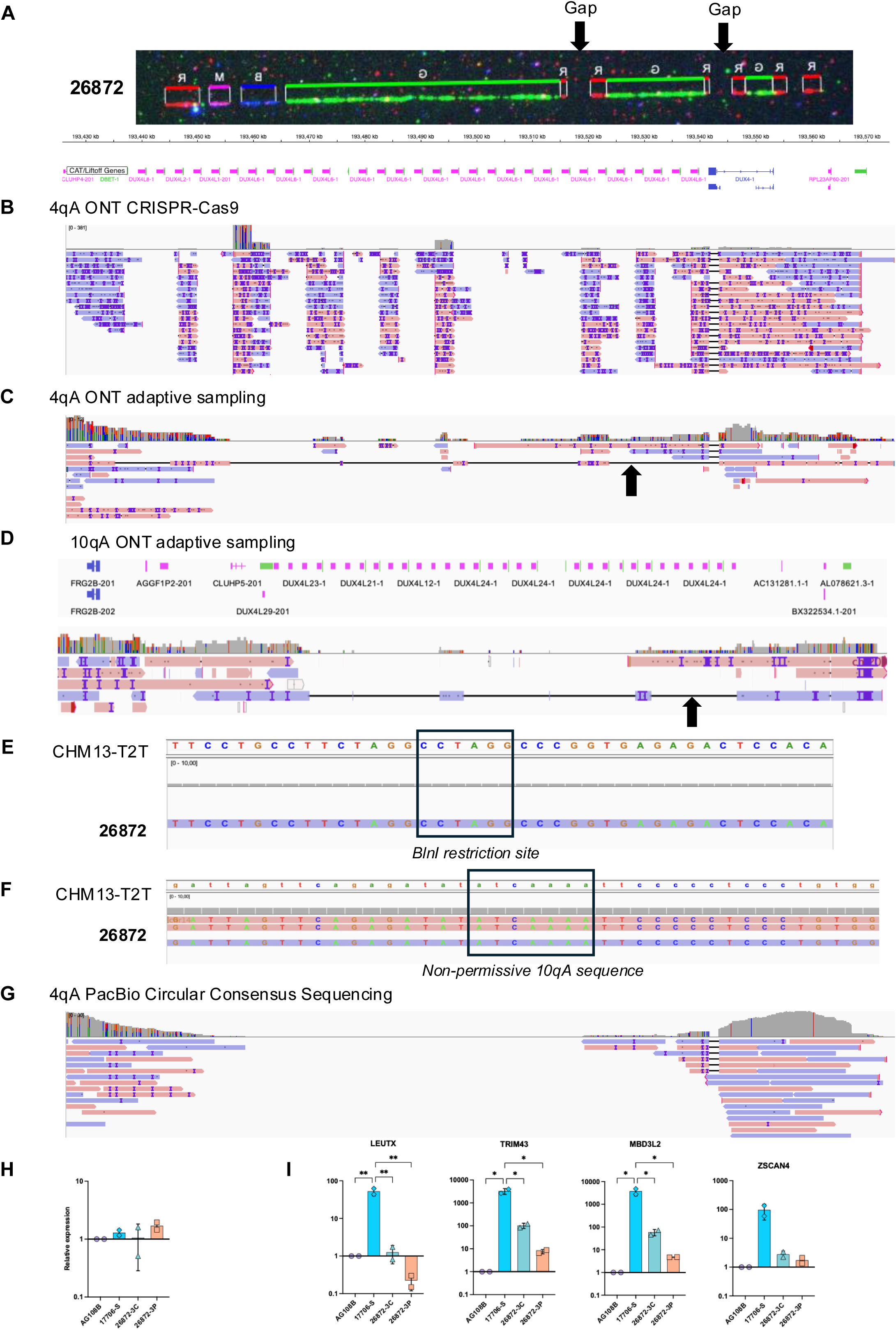
Comparison between ONT adaptive sampling and Pac Bio sequencing. **A**. Raw image from MC for patient 26872. From left to right: red probe specific to chromosome 4, pink and blue probes for the p13E11 region, green probe targeting D4Z4, red probe specific to the qA haplotype, a gap, and a red probe for the subtelomeric region, followed by two additional arrays showing the same probe order. **B.** A CRISPR-Cas9–based enrichment strategy was used, after a targeted cleavage guided by a gRNA specific to the D4Z4 array. Only targeted cuts guided by the D4Z4-specific gRNA are visualized using IGV. From top to bottom: position on the CHM13-T2T reference, RefSeq gene annotation, read coverage (0 to 381 reads), and aligned sequences. **C.** ONT Adaptive sampling analysis showing a long read (black arrow) spanning the entire 4q35 region from the p13E11 probe to the telomere. Tracks from top to bottom: read coverage (0 to 12 reads), and read alignment. **D.** IGV view of the T2T genome annotation (top track) and patient’s read alignment (bottom). A single long read spanning the entire 10qA locus and the 6 D4Z4 RUS is indicated by a black arrow. **E.** IGV view of chr10:134,659,678–134,659,717, showing the presence of a CCTAGG motif corresponding to the *Bln*I restriction site in both the reference sequence and the patient’s genome. **F.** IGV view of the chr10:134,726,525–134,726,564 position, showing the presence of a non-permissive pLAM sequence (ATCAAA) on the 10qA allele with 6 D4Z4 repeat units followed by a non-permissive A-type haplotype (black box). **G**. PacBio Circular Consensus Sequencing analysis (CCS), showing high-quality reads that do not fully span the D4Z4 array. From top to bottom: read coverage (0 to 30 reads), and aligned sequences. **H**. Expression of *DUX4* in the patient’s hiPSC-derived muscle tissue compared to hiPSC-derived muscle tissue for a healthy donor (AG108B) and FSHD1 patient (17706S) used as negative and positive controls, respectively. **F.** Expression of selected DUX4 target genes (*LEUTX* (Leucine Twenty homeobox), *TRIM43* (Tripartite Motif containing 43), *MBD3L2* (methyl CpG binding domain protein 3 L2) and *ZSCAN4* (Zinc finger and SCAN domain containing protein 4) in the same samples. **I.H.** Histogram bars display the results of two biological and three technical replicates for each sample. Two clones were analyzed for the patient. Statistical significance was determined using a Kruskal-Wallis statistical test. * p-value<0.05, ** p-value<0.005.

In addition, Adaptive Sampling provided one read of 166,523bp spanning chr4:193L410L529-193,574,945 and covering the entire 5RU 4qA sequence between the p13E11 locus and the distal pLAM as a continuous sequence without fragmentation (**Figure 3C**). Unfortunately, this approach still fails to capture the whole region and the proximal largest arrays (39 and 14 RUs). Nevertheless, the significantly higher number of reads generated (2.65M vs. 60.08k) compensates for the lower on-target percentage, ensuring sufficient coverage (**Table S1**).

By adaptive sampling, we also captured the entire 10qA short allele (6UR), with a single 70 kb read spanning the entire region (**Figure 3D**). On this 10qA read, six *Bln*I restriction sites were detected, supporting the absence of a 4q-10q rearrangements (**Figure 3E**). We confirmed the presence of an A-haplotype, carrying an ATCAAA sequence as expected for a 10qA allele instead of the ATTAAA pLAM element found in 4qA alleles (**Figure 3F**). The absence of a permissive polyadenylation signal confirms that the 10qA allele is not pathogenic in this patient.

Overall, Adaptive Sampling provided a more flexible approach, offering a broader and uninterrupted view of the regions of interest, even though only a single read covering the entire region of interest was obtained whereas CRISPR introduces targeted breaks that limit downstream analyses. Moreover, the improvement in sequencing quality observed with the R10 flow cells, coupled with the concurrent enhancement of base calling accuracy through the Dorado algorithm increased mapping quality.

### Side by side evaluation of ONT and PacBio genome sequencing for 4q35 structural variants detection

We next evaluated the performance of PacBio genome sequencing for patient 26872 (**Figure 3G**). Briefly, 9 µg high-molecular-weight genomic DNA was fragmented to ∼30 kb. Library preparation was performed using the SMRTbell® Express Template Prep Kit 2.0 (Pacific Biosciences). Sequencing was performed on a PacBio Revio system using SMRT Cell. Basecalling of sequenced bases and methylated bases was managed using SMRT Link v13.1.0.221970. High-fidelity (HiFi) reads were generated using the Circular Consensus Sequencing (CCS) mode with default parameters. Reads were aligned to the T2T-CHM13 v2.0 reference genome using pbmm2 (v1.9.0) with the --preset CCS option. The sequencing run provided a total of 87.76 Gb of data. We obtained a total of 5×10^6^ high-fidelity reads with 93.2% of them having a base quality of ≥Q30, ensuring good mapping accuracy and a total average read length of 17kb. Although the sequencing quality surpasses that of Nanopore and enables SNV analysis, the limitation of read length to 30 kb presents a significant drawback in the context of FSHD locus analysis. The longest reads obtained with PacBio only covered 7UR, while in adaptive sampling, we were able to obtain reads covering the entire locus (**Figure 3C**). An alternative to increase read depth at a specific locus involves following Nanopore’s recommendation of loading the equivalent of three libraries per run. However, this strategy demands a substantial amount of DNA and significantly raises the overall cost.

To further investigate the pathogenicity of the rearranged allele for this patient, we derived induced pluripotent stem cells (hiPSC) and produced HiPSC-derived muscle tissue using our previously described conditions (44) to analyze expression of *DUX4* and DUX4 target genes. *DUX4* expression is not detectable in control cells and slightly expressed in FSHD cells (17706S) or in muscle derived from two independent clones for patient 26872 (**Figure 3H**). We also detected the expression of 4 selected DUX4 target gene for this patient (**Figure 3H).** Differences in gene expression are not significantly increased compared to the healthy donor but significantly lower compared to FSHD cells, questioning the pathogenicity of the cis-triplication for this patient.

### All-in-One Genomic Profiling of 4q35 Using Nanopore Whole-Genome Sequencing

Considering the limitations of previous strategies for characterization of structural variants at the 4q35 locus, we next decided to evaluate the performance of ONT whole genome sequencing. To overcome the challenge of insufficient enrichment at the locus, we employed a PromethION Flow Cell, which offers enhanced performance. We sequenced a third patient carrying a 4qA 22+5RU *cis*-duplication. (27279, **Figure 4A**). The second 4q allele is a 4qA with 69 RUs. He also carries two 10qA with 22RU, as determined by MC (34). FSHD was part of a differential diagnosis for this patient, with a mild muscular weakness and essential tremor.

**Figure 4.**
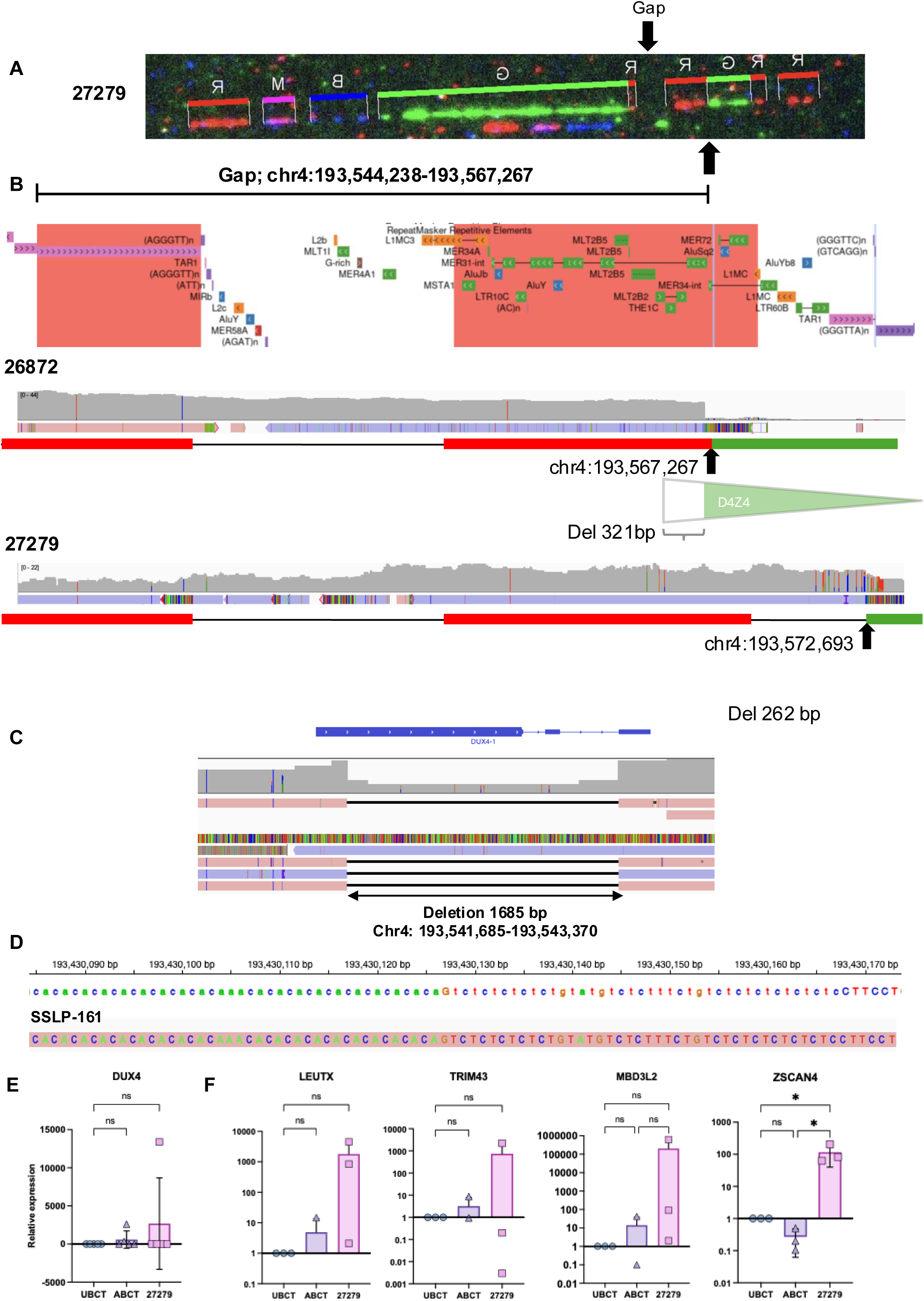
LSau-dependent D4Z4 array duplication leads to deletions of different sizes. **A.** Raw MC image for patient 27279. From left to right: the proximal red probe is specific to chromosome 4. The pink probe maps the proximal region that is shared between chromosomes 4 and 10. The blue probe maps to the p13E11 region. The green probe corresponds to D4Z4. Downstream of the green probe, the red probe corresponds to the qA haplotype. The first D4Z4 array (22 RUs) is followed by a second array that contains 5 RUs. The most distal red probe corresponds to the distal part of the subtelomeric region. **B.** Comparison of breakpoints between patients 26872 and 27279. From top to bottom: position of the “gap” on chr4 (CHM13 T2T; Chr4:193,544,238-193,567,267), repeat masker track from UCSC, coverage, aligned read in IGV, molecular combing probe for reference (qA probe and subtelomeric probe). Visualization using IGV of the breakpoint leading to deletion of a 262 bp fragment in the proximal part of D4Z4 and the *cis*-duplication at chr4:193,572,693 for patient 27279. Data were compared to the breakpoint of 321 bp at position chr4:193,567,267 identified for patient 26872. Breakpoints are indicated by black arrows. **C.** Close-up view of the *DUX4* sequence. For patient 27279, the 4qA-L haplotype includes a longer final intron, represented as a 1685 bp misalignment compared to the T2T reference genome (Chr4: 193,541,685-193,543,370) **D.** IGV view of the proximal polymorphic SSLP region spanning chr4:193,430,085-193,430,173. Upper panel, T2T CHM13 reference sequence. lower panel, patient showing a SSLP-161 (CA10 AA CA10 GT CT5 GT AT GT CT2 TT CT GT CT6) allele. **E.** Expression of *DUX4* in the patient’s muscle biopsy compared to control (12UBCT) and FSHD1 (12ABCT) myotubes obtained from hTERT-immortalized myoblasts and used as negative and positive controls, respectively. **F.** Expression of selected DUX4 target genes ((*LEUTX* (Leucine Twenty homeobox*), TRIM43* (Tripartite Motif containing 43), *MBD3L2* (methyl CpG binding domain protein 3 L2), *ZSCAN4* (Zinc finger and SCAN domain containing protein 4)) in the same samples. **E.F.** Quantitative PCR (qPCR) analysis was performed in triplicate using *HPRT* and *PPIA* as housekeeping genes. Relative expression values are presented in log₁₀ scale as mean ± SEM. Statistical analysis was performed using ANOVA,Tukey’s multiple comparisons test. A p-value < 0.05 was considered statistically significant (*).

Nine µg of DNA extracted from a muscular biopsy was used to prepare 3 libraries which were sequentially loaded with washes performed between each load on the FLO-PRO114M flow cell. Sequencing yielded 1.34 x 10^6^ reads, with an N50 of 7 kb, a maximum length of 520 kb, providing an overall satisfactory sequencing depth, quality and accuracy, considering that the DNA extraction was performed on a small muscle biopsy sample (**Table S1**).

In this patient, the *cis*-duplicated sequence lies in the same orientation and originates from chromosome 4, as evidenced by the presence of two *Xap*I restriction sites located +1535 and +4838 in the soft clipped sequence and the absence of *Bln*I restriction sites. These results support the hypothesis of an intrachromosomal rearrangement leading to *cis*-duplications (**Table S7, Figure 4B**). However, the breakpoint responsible for the *cis*-duplication differs from those observed in the other two patients and is positioned more distally, 5426 bp (chr4:193,567,267) further downstream of the breakpoint identified in patients 26872, 27858 and in the 2 patients reported previously (40) and downstream of the TAR (Telomere Associated Repeat) at chr4:193,572,693 (T2T-CHM13) (**Figure 4B**).

The first D4Z4 unit of the *cis*-duplicated array is deleted of 262 bp at its 5’ end (chr4:193,434,245-193,434,506 on the T2T). This deletion is smaller in size compared to the 321 bp deletion seen in the 2 other patients and reported elsewhere (40). Overall, this provides evidence that complex rearrangements do not originate from a common ancestor, but rather from homologous recombination between LSau repeat elements (βsatellite) contained within the D4Z4 unit and possibly in the distal 4qA region. A careful reexamination of the MC images indeed suggests that the breakpoint might not be the same. The subtelomeric probe following the gap is longer in patient 27279 (**Figure 4A**) compared to patient 26872 (**Figure 1B**). In patient 26872, an orange fusion signal is also observed between the red subtelomeric probe and the green D4Z4 probe (**Figure 1B, orange arrow**), which is not seen in patient 27279. Because of the low resolution of MC, this result that was initially overlooked is now explained by ONT results that shows that the breakpoint is not located within the subtelomeric probe region.

For this patient, we successfully sequenced the gap between the pLAM region and the telomeric T_2_AG_3_ repeats. The sequence shows 99.2% identity with the CHM13-T2T 4q reference and 98.7% with chromosome 10q. The patient also carries at least one 4qA-L allele observed as the corresponding 1,685 bp deletion when aligned to the CHM13-T2T reference genome (**Figure 4C**). We also analyzed the proximal SSLP microsatellite (chr4:193,430,085–193,430,167, CHM13-T2T) and found that the patient carries an SSLP-161 allele (**Figure 4D**).

Having access to a muscle biopsy for this patient, we next tested for expression of *DUX4* and selected DUX4 target genes by RT-qPCR in comparison to myotubes obtained from hTERT-immortalized muscle cell lines (control and FSHD1). Detection of *DUX4* was variable in the biopsy sample (**Figure 4E**) together with expression of different DUX4 target genes, except for *ZSCAN4* (**Figure 4F**) suggesting that for this patient, the rearrangement might be associated with molecular features of the disease but analysis of the muscle biopsy was not informative in that case.

### Nanopore Sequencing Provides Critical Insights Beyond Optical Genome Mapping in Complex Genotype Cases

Using the same approach, we next reanalyzed the genotype of four patients affected with FSHD and the segregation of FSHD-associated genetic features across generations. In this family, individuals 17370, 15674 and 16705 were previously reported as carrying a *cis*-duplication on chromosome 4 identified by MC(35) (50+5 RUs; **Figure 5A**) together with a pathogenic variant in *SMCHD1* (Exon 28, c.3631C>T; p.(Gln1211*)) and D4Z4 hypomethylation (23-27%). The index case (15674) also carries a 4qB allele (16 RUs) and a short 10qA allele (6 RUs) (**Figure 5B**), raising questions whether the *cis*-duplicated allele is pathogenic and permissive for *DUX4* expression.

**Figure 5.**
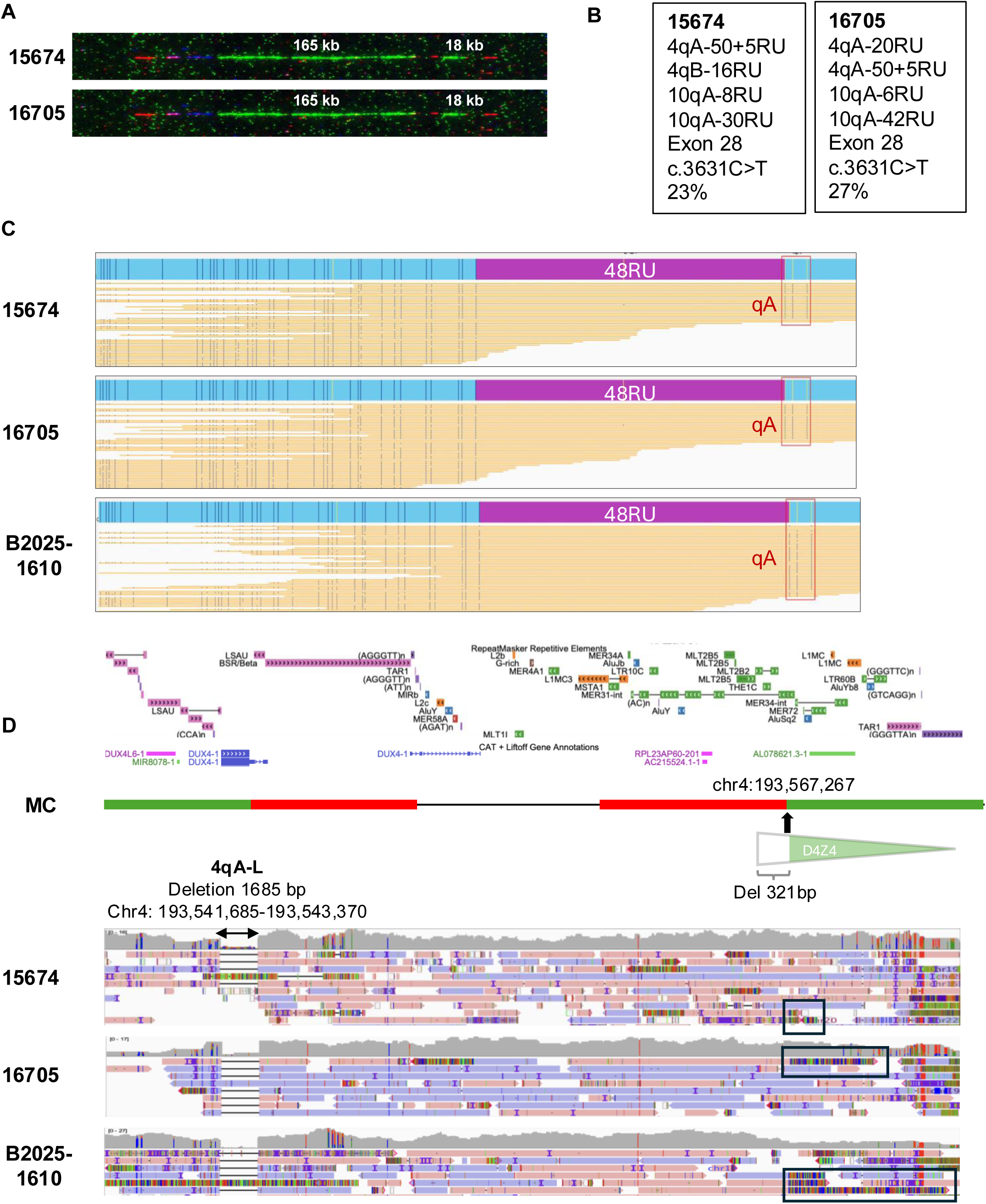
Nanopore sequencing complements Bionano Optical Genome Mapping in case of complex genotypes. **A.** Raw MC images for patients 15674 and 16705. From left to right: the proximal red probe is specific to chromosome 4. The pink probe maps the proximal region that is shared between chromosomes 4 and 10. The blue probe maps to the p13E11 region. The green probe corresponds to D4Z4. Downstream of the green probe, the red probe corresponds to the qA haplotype. The first D4Z4 array (50 RUs, 165 kb) is followed by a second array that contains 5 RUs (18 kb). The most distal red probe corresponds to the distal part of the subtelomeric region. **B.** Genotype for patients 15674; 16705 as previously reported (35). In addition to the *cis*-duplication, they all carry a pathogenic variant in *SMCHD1* (Exon 28; c.3631C>T; p.(Gln1211*)). **C.** Bionano Optical Genome Mapping followed by FSHD EnFocus analysis of the 4q35 region for patients 15674 and 16705. Patient B2025-1610 is the sister of 15674 and was more recently investigated, only by OGM. The D4Z4 array is represented in purple. For all three patients, the *cis*-duplication was missed by the FSHD EnFocus tool and the 4qA allele was estimated to be 48 RUs in size. The *cis*-duplication was visualized by the presence of an atypical incomplete A-haplotype pattern downstream of the D4Z4 array (red box). Patients 15674 and B2025-1610, also carries a 4qB allele estimated of 15 RUs. Patient B2025-1610 carries a 10qA at 8 RUs and one at 40 RUs. **D**. Visualization using IGV (Integrated Genome Viewer) of Oxford nanopore sequencing data for patients 15674, 16705 and B2025-1610. From top to bottom: RepeatMasker Track, RefSeq track, corresponding MC image, read coverage (ranging from 0 to 16-17-27 reads for 15674, 16705 and B2025-1610 respectively), and aligned sequences. The Black box indicates the split reads corresponding to the breakpoint located at chr4:193,567,267. The 4qA-L allele is visible as a 1685bp deletion (Chr4: 193,541,685-193,543,370).

More recently, her sibling (B2025-1610) was clinically diagnosed with FSHD. As MC is no longer available, molecular diagnosis for this patient was performed only by OGM and compared to the analysis of her relatives (**Figure 5C**). The *cis*-duplicated allele was not detected by the FSHD Enfocus analysis tool that reported the same allele containing 48 RUs but the presence of an atypical distal pattern (4qA haplotype) requiring manual curation for all three patients (**Figure 5C**). Patient B2025-1610 also carries a 4qB allele (15 RUs) and short 10qA allele (8 RUs).

We subsequently performed ONT whole-genome sequencing on all three patients to confirm whether the *cis*-duplicated allele is permissive for *DUX4* expression and exhibits the classical genetic features associated with the disease (**Figure 5D**). For each patient, nine µg of DNA extracted from whole blood was used to prepare 3 libraries, which were sequentially loaded with washes performed between each load on the FLO-PRO114M flow cell. Sequencing yielded between 8 and 19 million reads per patient, with a total yield ranging from 34 to 83 Gb. The N50 read length varied from 8.8 kb to 11 kb, and the maximum read length reached up to 1 Mb. Specifically, patient 15674 generated 17 million reads for a total of 53 Gb, with an N50 of 8.8 kb and a maximum read length of 1 Mb; patient 16705 generated 8 million reads (34 Gb), with an N50 of 11 kb and a maximum read length of 761 kb; and patient B2025-1610 generated 19 million reads (83 Gb), with an N50 of 10 kb and a maximum read length of 614 kb (**Table S2**).

We successfully sequenced the “gap” sequence that shows 99.5%, 99.4%, 99.0% identity with the CHM13-T2T 4q reference and 98.8%, 98.8%, 98.5% with the 10q reference for patients 15674, 16705, and B2025-1610, respectively. The 3 patients show the recurrent deletion of 321 bp located in the proximal part of each *cis*-duplicated array (indicated by split reads, black boxes, **Figure 5D**). The distal breakpoint located at chr4:193,567,267 is identical to patients 27858 who carries a *cis*-duplication or patient 26872 carrying a *cis*-triplication. We observed the presence of a homozygous 4qA-L haplotype on chromosome 4 for patients 15674 and B2025-1610, consistent with the other allele being 4qB. For the mother (16705), two out of fourteen reads were 4q-S type, corresponding to the haplotype of the other 4qA at 20RUs (**Figure 5D, Figure S5A**). We also confirmed the presence of the *SMCHD1* variant for all three patients.

For each individual, we confirmed the A-haplotype on chromosome 10 containing a 10q-specific pLAM corresponding to a ATCAAA sequence instead of the ATTAAA pLAM element specific to 4qA alleles (**Figure S5B**). The absence of a permissive polyadenylation signal on the 10qA containing 8RUs confirms that the 10qA allele is not pathogenic in this family. We report the presence of *Xap*I restriction sites within the *cis*-duplicated arrays for all three cases, supporting a 4q origin of the *cis*-duplication (**Table S7**). In patient 16705, *Xap*I sites were detected at position +1484 and within the duplicated segment. The +1484 *Xap*I site and another at +4768 were also present in patient B2025-1610. For patient 15674, soft-clipped sequences at the breakpoint were too short (427 bp) to confidently assess the presence of these restriction sites (**Figure 5D**). We also analyzed the proximal SSLP microsatellite (chr4:193,430,085–193,430,167, CHM13-T2T) and found that all patients carry a homozygous SSLP-161 allele (Figure S5C). For this patient, all of these findings support the pathogenicity of the cis-duplication.

### 10q *cis*-duplication supports the hypothesis that breakpoints are not recurrent but occur randomly as a result of subtelomeric instability

We next analyzed patient 240080 who presents with a clinical phenotype highly evocative of FSHD and classified as FSHD2. He carries two 4qA alleles, one with 11 RU and one estimated at 45 RU. He also carries a heterozygous *SMCHD1* variant (NM_015295.3:c.1295T>C:p.(Phe432Ser)) located within the ATPase domain. D4Z4 DNA methylation level at DR1 level in blood DNA was estimated around 35% by bisulfite sequencing, consistent with a diagnosis of FSHD2 (45). He also carries two 10qA alleles, one 10qA with 37 RUs, and one 10qA with a *cis*-duplication estimated at 42+11RU (first D4Z4 array of 140 kb and second array of 35 kb) as determined by MC (**Figure 6A**).

**Figure 6.**
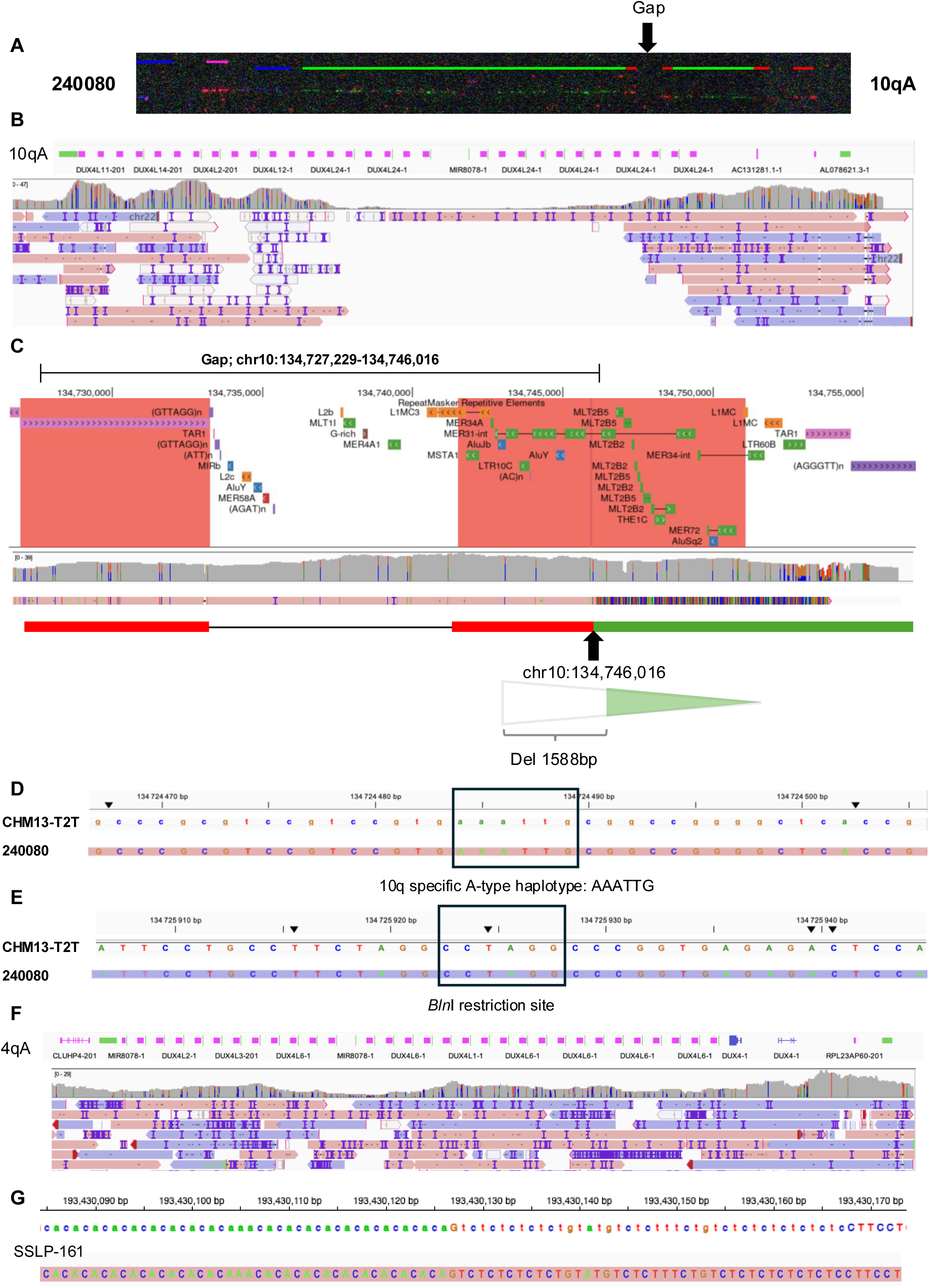
Characterization of a *cis*-duplicated 10qA allele. **A.** Raw MC image of the 10q locus in patient 240080. From left to right: the proximal blue probe is specific to chromosome 10. The pink probe maps to the proximal region shared by chromosomes 4 and 10. The blue probe maps to p13E11. The green probe corresponds to D4Z4. Downstream of the green probe, the red probe corresponds to the qA haplotype. Two array of 11 RUs, with a region that is not covered by MC probes (gap). **B.** Visualization of 10q ONT sequencing reads aligned to the CHM13-T2T reference genome using IGV. From top to bottom: position on the CHM13-T2T reference, RefSeq gene annotation, read coverage (0 to 47 reads), and aligned sequences. **C.** Close-up view of the breakpoint at chr10:134,727,229-134,746,016 located within the MER31-int element, 880 bp downstream of an AluY element. This duplicated MER31-int is oriented head-to-tail and shows a 1588 bp deletion in 5′ of the first D4Z4 unit. **D.** Sequence analysis of the last D4Z4 repeat showing the absence of *Xa*pI restriction site at chr10:134,724,484–134,724,489 and the presence of a distal AAATTG instead of the 4qA AAATTC PAS sequence. **E.** Sequence analysis of the last D4Z4 repeat shows presence of a *Bln*I-permissive site (CCTAGG) at chr10:134,725,922– 134,725,927, supporting the absence of a 4q–10q translocation. **F.** Sequencing of the 4qA allele did not yield long reads covering the entire 11 RU array. From top to bottom: position on the CHM13-T2T reference, RefSeq gene annotation, read coverage (0 to 29 reads), and aligned sequences. **G.** IGV view of the SSLP polymorphism at the 4qA locus shows a homozygous SSLP-161 allele (CA10 AA CA10 GT CT5 GT AT GT CT2 TT CT GT CT6).

Whole-genome nanopore sequencing was performed using a PromethION flow cell (FLO-PRO114M). A total of 9 µg of DNA extracted from frozen blood was used for preparation of three library, sequentially loaded with intermediate washes. The run yielded 6.6 million reads and 57LGb of data, with an N50 of 19.5 kb, a maximum read length of 1LMb. This sequencing provided a good depth and satisfactory read accuracy, confirming that whole-genome nanopore sequencing on a PromethION flow cell is an efficient approach for analysis of the FSHD locus. We were not able to sequence the entire 10q duplicated region containing 42+11RUs as the longest reads spanned only the five most distal D4Z4 units but also the entire downstream sequence up to the telomere (**Table S1, Figure 6B**).

At 10qA, the “gap” region that is not covered by MC probes was fully resolved and shows 99.3% homology with the telomeric sequence of the CHM13-T2T assembly (chr10:134,727,229-134,746,016 (18Mb)), and 98.7% homology with the 4q sequence (NCBI Blast). The breakpoint is located at chr10:134,746,016 (CHM13-T2T), within the region mapped by the red subtelomeric probe, in the MER31-int element, 880 bp downstream of an AluY element (**Figure 6C**). This MER31-int element is duplicated *in cis* in a head-to-tail orientation, leading to a deletion of 1588 bp in the 5′ end of the first D4Z4 element. The rearrangement originates from chromosome 10, as evidenced by the presence of *Bln*I restriction sites in soft-clipped reads at positions +1656, +4962, and +8261 (**Table S7**). At chr10:134,724,484–134,724,489, we consistently observed the presence of a AAATTG sequence (*Xap*I site: AAATTC) (**Figure 6D**), and a conserved CCTAGG sequence at chr10:134,725,922–134,725,927, that corresponds to a *Bln*I restriction site characteristic of 10q D4Z4 (**Figure 6E**). This result further supports the absence of a 4q–10q translocation event associated with *cis*-duplications. We also sequenced the 4qA allele but could not obtain reads of the entire 11RUs array (**Figure 6F**). The patient carries a homozygous SSLP 161 at the 4qA locus (**Figure 6G**), supporting the possible association between this SSLP and instability of the locus.

### D4Z4 methylation analysis using long read sequencing as an alternative to bisulfite sequencing

Following ONT long read sequencing, we performed methylation plotting across the whole 4q35 locus using methylartist. version 1.5 (**Figure 7A, Figure S6**), the last D4Z4 array (**Figure 7B, Figure S7;8**) and pLAM region (**Figure 7C**, **Figure S9**) for the different patients. Methylartist processes BAM files containing modification tags (MM/ML), enabling visualization of methylation patterns at a single-molecule resolution, as well as aggregated methylation frequencies across the targeted genomic locus, focusing on CpG motifs. For patient 240080 who carries a variant in *SMCHD1*, the methylated haplotype at the chr4:193,429,551-193,574,945 locus exhibits a cyclical methylation profile with peaks of methylation approximately every 3.3 kb, matching individual D4Z4 repeat units. Mean methylation starts around 25% for the most proximal D4Z4 repeat and progressively increases over the first 10 units, approaching 50% of methylated CGs in the most distal D4Z4 unit (**Figure 7A**). However, Interpretation should consider potential biases of coverage, methylation calling and allele phasing. For this patient, analysis of the last D4Z4 unit and the junction with the pLAM region (chr4:193,539,766–193,543,060) revealed a marked hypomethylation of the LSAU and COMP-subunit-LSAU-BSAT elements (chr4:193,539,766– 193,540,742) (**Figure 7B, Sup Figure 8D**). This region encompasses the first 977 bp of the D4Z4 unit and includes the DR1 region (positions 556 to 808 bp within D4Z4). In this patient, bisulfite sequencing, provided an average methylation level across all DR1 sites (4q and 10q) locus of 35% methylation consistent with the presence of a pathogenic *SMCHD1* variant(45). The Methylartist profile reveals consistently high methylation levels across the pLAM region which has been suggested to display FSHD2-associated hypomethylation(46), remaining above 50% for all patients analyzed, which does not support fully support its discriminative value as diagnostic marker for ONT sequencing (**Figure 7C, Figure S9**).

**Figure 7.**
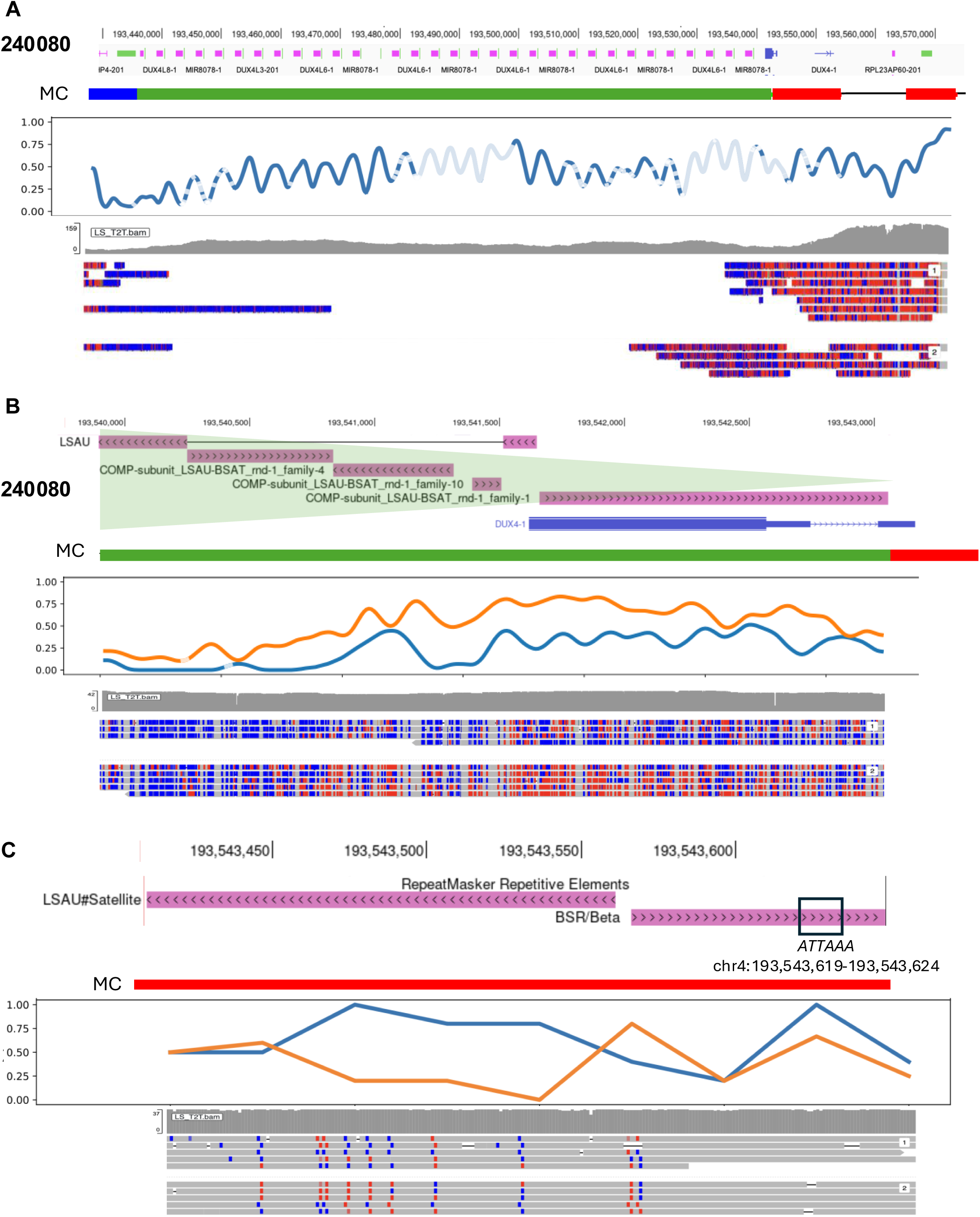
Methylation analysis by ONT sequencing as an alternative to bisulfite sequencing. **A.** Methylation profile across the entire 4q35 locus (chr4:193,429,551–193,574,945; CHM13-T2T) in patient 240080, obtained using methylartist v1.5. From top to bottom: RefSeq track from UCSC, position of the MC probes (from left to right, p13E11 (Blue), D4Z4 (green), A-type haplotype (red) and distal telomeric probe (red)), methylartist methylation plot (*x-axis*: chromosomal position, *y axis*: mean methylation percentage). The different shades of blue indicate the coverage depth (from 0 to 159). Sequences were aligned using IGV with the “base modification 2 color” parameter. **B.** Focus on the last D4Z4 repeat unit and its junction with the pLAM (chr4:193,539,766–193,543,060, CHM13-T2T) for patient 240080. From top to bottom: RefSeq track from UCSC, position of the MC probes (from left to right, D4Z4 (green), A-type haplotype (red)), methylartist methylation plot with haplotype 1 indicated in blue, and haplotype 2 in orange (*x-axis*: chromosomal position, *y-axis*: mean methylation percentage), depth from 0-42 and aligned sequences. **C.** Methylation pattern of the pLAM region (chr4: 193543409-193543649, CHM13-T2T). From top to bottom: RefSeq track from UCSC, position of the red MC probe corresponding to the A-type haplotype, methylartist methylation plot with haplotype 1 indicated in orange, haplotype 2 in blue (*x-axis*: chromosomal position, *y-axis*: mean methylation percentage), coverage from 0-37 and aligned sequences.

We next performed methylation analysis using modkit with the pileup command to extract CpG methylation levels for Nanopore and PacBio reads. Basecalling and alignment were performed with Dorado v 0.9.1 for ONT and SMRT Link v13.1 for PacBio, which integrates modified base detection to assign methylation probabilities directly from raw reads. By default, a filtering threshold of 0.7 was applied for cytosine methylation. This generated a pileup BED file containing, for each CpG site, the percentage of methylation (5mC, labeled as m).

For each patient, we first compared the long-read methylation at DR1 for the most distal D4Z4 unit extracted using modkit with standard bisulfite sequencing (**Table S2, Figure S8A,B;**). A custom Python script was used to filter these data for the predefined DR1 (9, 47) (**Figure S8B)** and pLAM regions (46, 48) **(Figure S9)**, to compute average modification levels, and generate bar plots showing the methylation across the sequences of interest.

Overall, we observed that mean methylation values derived from Nanopore or PacBio data tended to be different than those obtained by sodium bisulfite sequencing, with the most pronounced discrepancies observed for CRISPR-enriched Nanopore runs. We attributed these differences to the limited coverage, read depth and low resolution. In patient 26872, bisulfite sequencing showed 48.45% methylation at DR1, whereas CRISPR-enriched Nanopore reads gave 66.2%. For adaptive sampling sequencing the methylation level was estimated around 84.9%, and 63.9% by PacBio data (**Figure S8B**). Due to the low read depth, we did not analyze patient 27858. For patient 27279, Nanopore genome-wide reads yielded 45.5% methylation versus 59.60% by sodium bisulfite sequencing. On the other hand, patients 240080, 15574 and 16705, sequenced on a PromethION using high-quality genomic DNA from whole blood, showed improved consistency between sodium bisulfite sequencing (35%, 23%, 27%, respectively) and Nanopore (23.4%, 16,7%, 39.9%), supporting the reliability of this most recent approach (**Figure 7B, S8D**). As observed using Methylartist, the level of methylation of the pLAM region is high (>50%) for all samples tested (**Figure S9**).

Altogether, these results highlight both the technical progress and the importance of input quality in accurately measuring methylation from long-read data. The performance and higher coverage observed with patient 240080 and achieved with WGS suggests that with high-quality samples and optimal sequencing strategies, long-read methylation profiling could become a robust alternative for assessing locus-specific methylation, including in clinically relevant regions like D4Z4. Validation across different threshold settings and validation on more patients, including FSHD1 and FSHD2 patients is required to ensure a more accurate and consistent interpretation of ONT methylation data.

## Discussion

Using Molecular Combing (MC), we identified the existence of complex rearrangements of 4q alleles in patients affected with FSHD (34–36). *Cis*-duplications that usually consist in the *in-cis* duplication of D4Z4 arrays of different size with a large proximal array (>11 D4Z4 units) followed by a contracted distal array (<25 kb) represents the most frequent category of these complex structural variants found at 4q and 10q loci and were reported by others since our initial publication (40, 49). *Cis*-duplicated arrays are separated by large regions of 20-40 kb that are not covered by MC probes (so-called “gap”).

With the goal of characterizing the molecular event associated with these rearrangements, we evaluated the performance of different long read sequencing strategies. Earlier results on Whole Genome Sequencing using the Nanopore technology on a MinION (32) or a GridION (50) sequencer revealed a low coverage at the 4q and 10q loci, emphasizing the need for a targeted approach to enrich the number of reads in these regions. Whole Genome Sequencing on PromethION showed improved coverage averaging 29X, but at a comparatively higher sequencing cost (51). As an alternative, targeted Cas9 sequencing using gRNA on Nanopore (nCATS) has been used to sequence the 4q35 region using multiple guides flanking the D4Z4 (32, 40, 42, 43, 50, 51). Compared to earlier runs on MinION with CRISPR sequencing, whole genome Promethion provides longer and higher-quality reads with significantly improved depth, which enhances structural variant detection and methylation analysis. In our hands, sequencing from high-quality blood DNA on PromethION resulted in the most consistent and reliable dataset (e.g. patient 240080; 16705; 15674; B2025-1610), suggesting that both DNA source and platform choice are critical for accurate characterization of the 4q35 and 10q26 loci.

We demonstrate here that duplications involve intrachromosomal recombination between LSau/βsatellite elements. In several cases, the *cis*-rearrangements leads to the deletion of 321 bp in the 5’ end of the first D4Z4 unit, consistent with previous reports (40). The resulting deletion encompasses the proximal Lsau element within the D4Z4 repeat and β-satellite elements within the distal qA haplotype. Notably, we describe here a novel structural 4q35 variant involving a shorter proximal deletion of 262 bp resulting from recombination between Lsau and β-satellite sequences. These findings imply variability in the exact breakpoint positions, while still implicating the same repetitive elements. This suggests a specific structural context favoring a conserved yet flexible recombination-prone architecture at the 4q35 locus, modulated by the presence of satellite elements. Notably, all rearrangements characterized by ONT long-read sequencing are consistently associated with 4qA-type alleles and more particularly, 4qA-L haplotypes that are five times less common than the 4qA-S haplotype (37). This observation supports the notion that qA haplotypes, particularly those containing an extensive array of β-satellite repeats distal to D4Z4 (36), are more susceptible to recombination events and, consequently, more frequently associated with D4Z4 array contraction as in FSHD1. Supporting this hypothesis, β-satellite repeats are known for their high recombinogenic potential, which facilitates structural rearrangements due to meiotic misalignment in the human genome (52, 53). We further showed that rearrangements occurring at the 10q26 allele might involve recombination between other repetitive elements leading to a larger deletion within the most proximal D4Z4 element, as also reported for other patients(54).

Subtelomeres, are complex genomic regions at the transition between chromosome-specific sequences and terminal telomeric repeats extending over a distance of 8 to 300 kb (55). These regions of transition between chromosome-specific sequences and terminal telomeric microsatellites show a mosaic structure composed of homologous duplicons shared between several chromosomal ends. Subtelomeres encounter a higher frequency of chromosomal inter- and intra-chromosomal exchanges that occurs through mechanisms such as crossing-over, gene conversion, or non-homologous end joining (NHEJ). The 4q35 subtelomeric qA and qB haplotypes are composed of repeated elements and share a strong homology. Large repetitive arrays, such as those present at 4q35 and 10q26 loci are prone to structural variants and therefore remain partially assembled in the reference genome, including the most recent T2T assembly. For instance, the 10qB-type allele which is rare in general population is absent from all reference genomes (hg35, hg36, hg37, hg38) and the CHM13-T2T version 2.0 assembly only reports 4qA short alleles (4qS) (38). However, despite frequent recombination upstream of the D4Z4 array (56), the distinction between qA and qB is remarkably stable across generations, suggesting selective pressure to maintain both variants (57). The qA haplotype that is associated with FSHD (58) (57), contains downstream of the D4Z4 array, an array of β-satellite repeats (BSR), composed of ∼68 bp repeat units, that are absent in qB, suggesting a potential role for β-satellite elements in FSHD pathogenesis. We further showed here the presence of multiple families of microsatellites, G-rich sequences, and old transposable elements distal to the BSR (**Figure S4)** and corresponding to the so-called “gap” region that creates a permissive environment for rearrangements. Furthermore, downstream of the BSR, we noticed the presence of (AGGGTT)n interstitial telomeric repeats (ITS), which can be locally amplified through polymerase slippage and increase the risk of structural rearrangements such as fusions and inversions (59) (60). The region also contains numerous Long Terminal Repeats (LTRs, 12 hits) with homologous 5’ and 3’ LTRs, which makes them prone to recombination(61). Retrotransposons found in the “gap” region derive from ancient endogenous retroviruses, such as MER31-int or MLT2B5 (62). We also identified several SINEs (Short Interspersed Nuclear Elements, 6 hits in our locus) that are small non-autonomous retroelements (100– 300 bp) responsible for non-allelic homologous recombination (NAHR), often leading to deletions or duplications (63). In addition, some of these elements may also have regulatory functions like the THE1C that display enhancer activity in cancer (64), the MIR family (65), G-rich G-quadruplexes (G4) (such as GGGGxx motifs) or ITS (66) involved in transcriptional regulation (67) and double-strand breaks (68). Besides contributing to the instability of the locus, many of these elements might also contribute to the silencing of nearby genes through telomeric position-effect (TPE) (55) or in gene sllencing at variable distances from the telomere, as reported for *DUX4* or *SORBS2* (69, 70) at the 4q35 locus.

Patient 26872 carries a single 4qA allele in *cis*-triplication configuration, and a 4qB allele on the other chromosome. This patient is homozygous for the 4qA-L haplotype and SSLP161, which supports the hypothesis of an intrachromosomal rearrangement rather than an interchromosomal event. Patients 26872, 27279, 27858, 16705; 15674; B2025-1610 share the same identical 4q breakpoint at chr4:193,567,267, which was also described in two other examples (40). In contrast, patient 27279 presents a novel breakpoint at chr4:193,572,693, never reported before. Breakpoints at the 4q35 locus differ from the breakpoint identified for patient 240080 who carries an intra-chromosomal 10q *cis*-duplication in the absence of a 4q– 10q translocation and from the breakpoints identified elsewhere(54). In all cases, the rearranged 4q alleles segregate with the 4qA-L haplotype and SSLP161, suggesting these genomic features may represent a predisposition factor. However, this remains to be evaluated on a larger cohort. Given the abundance of repeated elements in this locus, such rearrangements likely arise independently in the general population rather than being inherited from a common ancestor.

A previous study proposed that *cis*-duplicated alleles might exert pathogenic effects in a dominant manner(40). However, methodological limitations might mitigate this interpretation. The assessment of D4Z4 methylation levels was based on the per-read percentage of methylated CpGs (mCG) in D4Z4-enriched reads for approximately 320 CpG per 3.3 kb read, lacking the positional resolution of known differentially methylated regions such as DR1(9, 47) or the *Fse*I restriction site(71). The pathogenic potential of the recombined allele was inferred using the Log2p * Log2d of proximal and distal D4Z4 repeat counts, respectively. However, methylation differences between the *cis*-duplicated arrays complicate this interpretation. For example, in patient Rf2988.202(40), methylation levels differed markedly between the proximal 2 RUs (19%) and the distal 10-RUs D4Z4 array (44%). The authors concluded that the proximal region showed methylation patterns consistent with typical FSHD1 alleles. However, this conclusion is undermined by the imbalance in the number of reads with approximately 200 reads analyzed for the proximal short array, compared to only 4 reads for the distal region of the duplicated allele. This disparity, is likely stemming from the CRISPR-based enrichment method, weakening the robustness of the conclusion that has to be considered with caution regarding a diagnosis context.

Overall, structural variants found in FSHD should be interpreted with caution. Asymptomatic or non-penetrant carriers of *cis*-duplicated alleles have also been identified in multiple families. (34–36, 40) and while individuals 27279 and 26872 do not exhibit a clear FSHD clinical phenotype, other patients (27858, 240080, 16705, 15674, and B2025-1610) have been clinically diagnosed with FSHD. In patients 27858, the *cis*-duplication represents the only known genetic alteration compatible with the diagnosis, in the absence of *SMCHD1* variants or a contracted 4qA allele while it segregates with other FSHD-associated features in other cases (15674, 16705, B2025-1610. Results obtained for patient 26872 using different approaches remain inconclusive.

## Conclusions

Our observations support the view that the 4q subtelomeric region, already recognized for its evolutionary plasticity, is highly susceptible to recent structural rearrangements. Moreover, they expand the known spectrum of structural variations at the 4q35 locus by identifying distinct *cis*-duplicated alleles with variable distal breakpoints, thereby challenging the prevailing view of a single ancestral, recurrent rearrangement pattern. Overall, caution is needed when interpreting the pathogenic significance of complex rearrangements occurring at the 4q35 locus, in the absence of other FSHD-associated features as not all rearrangements are pathogenic or directly linked to a classical FSHD phenotype

## Data Availability

All data produced in the present work are contained in the manuscript

## Ethics approval and consent to participate

Samples were provided by the Center for biological Resources (Department of Medical Genetics, Biogenopole, La Timone hospital) with the AC 2011-1312 and N°IE-2013-710 accreditation numbers. All individuals have provided written informed consent for the use of DNA sample for medical research and the study was done in accordance with the Declaration of Helsinki. All patients were diagnosed at the Department of Medical Genetics, La Timone Hospital Marseille either by Southern blotting or Molecular Combing.

## Consent for publication

I confirm that authors have approved the manuscript for submission and that the content of the manuscript has not been published, or submitted for publication elsewhere.

## Acknowledgements

We are indebted and thank all patients for participating in this study.

