## Supplementary figures for "Comparison of long-read sequencing strategies for resolving complex genotypes at Facioscapulohumeral dystrophy-associated loci"

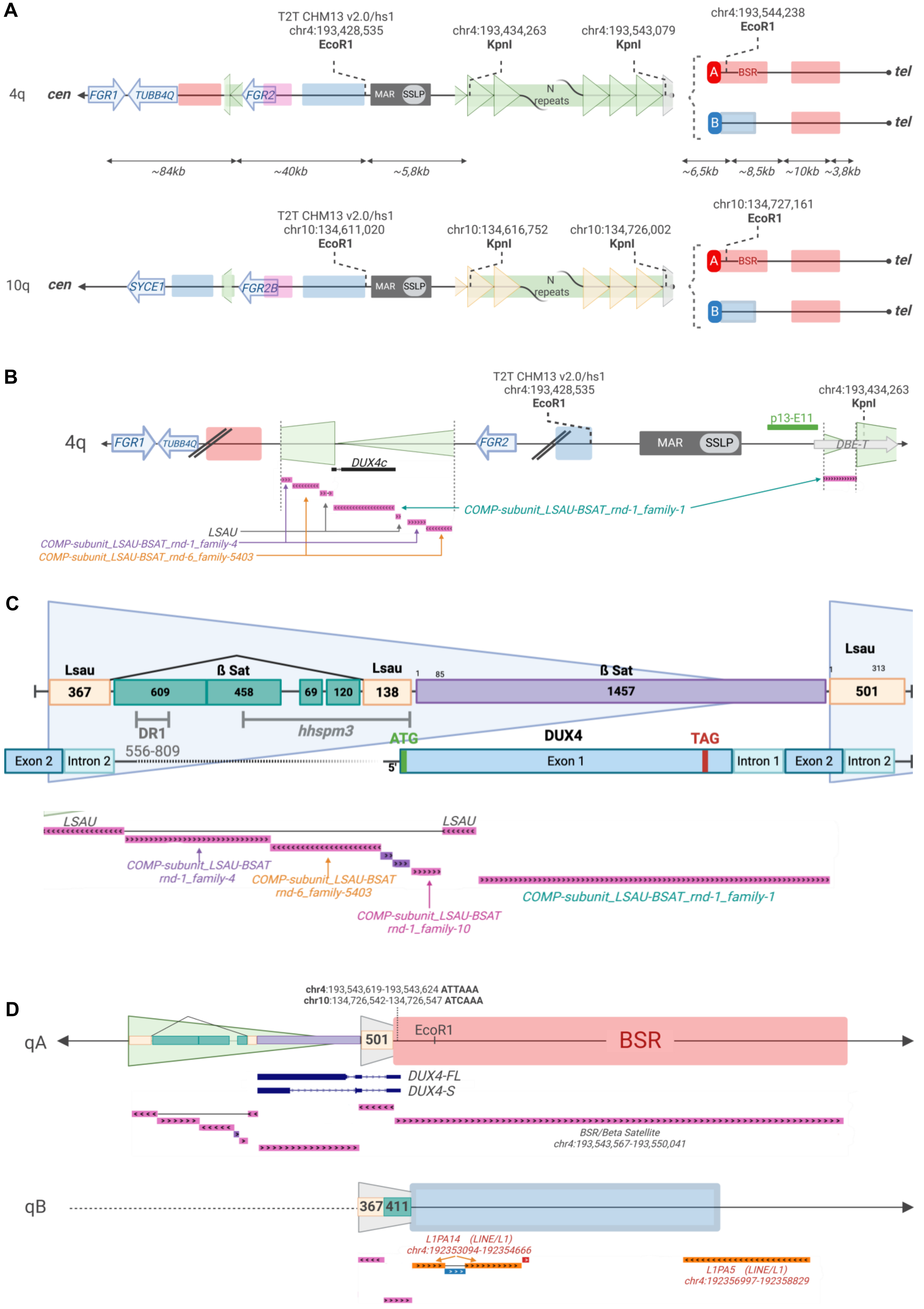

Tardy *et al*

**Supplementary figure 1**

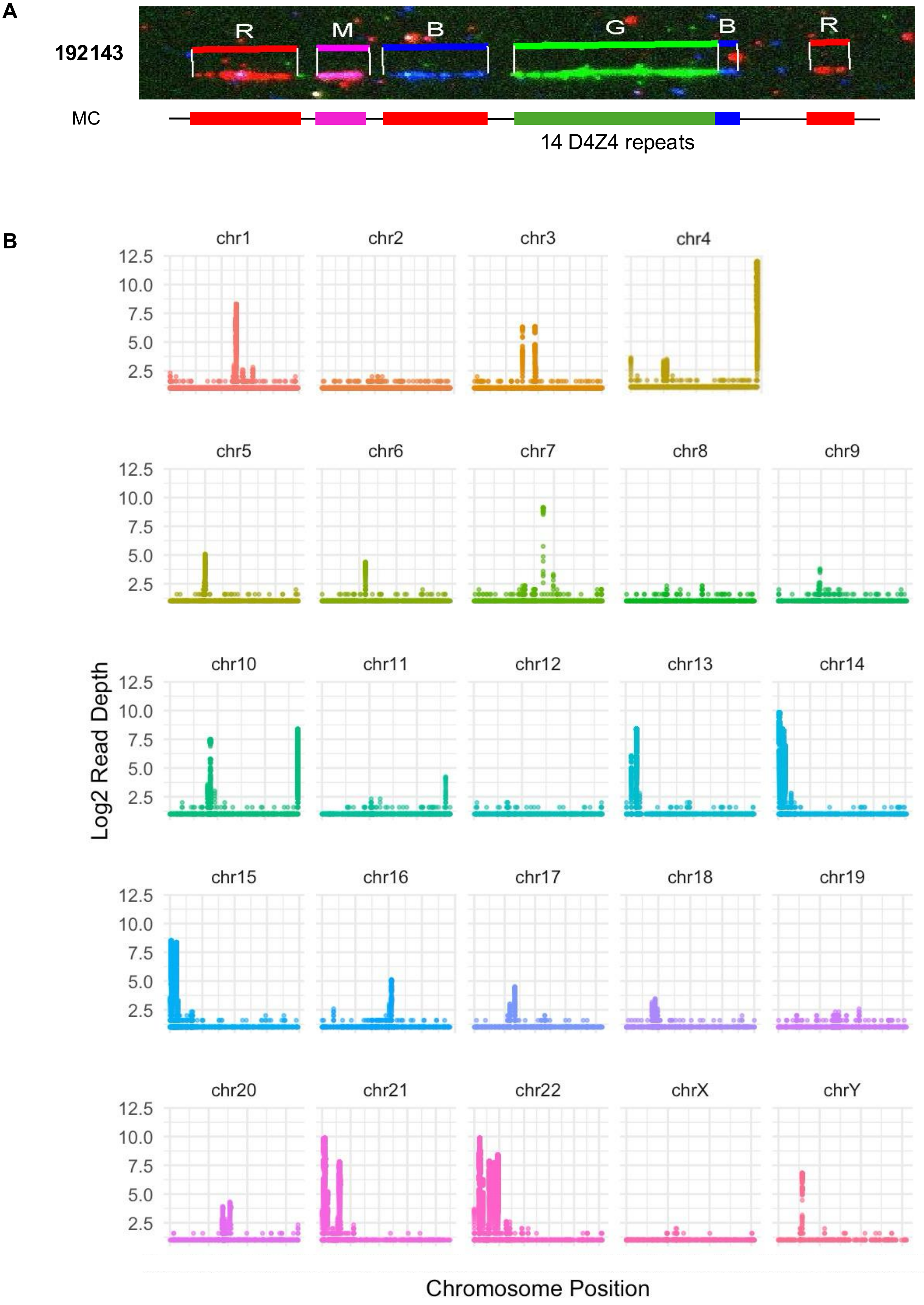

Tardy *et al* **Supplementary Figure 2**

A

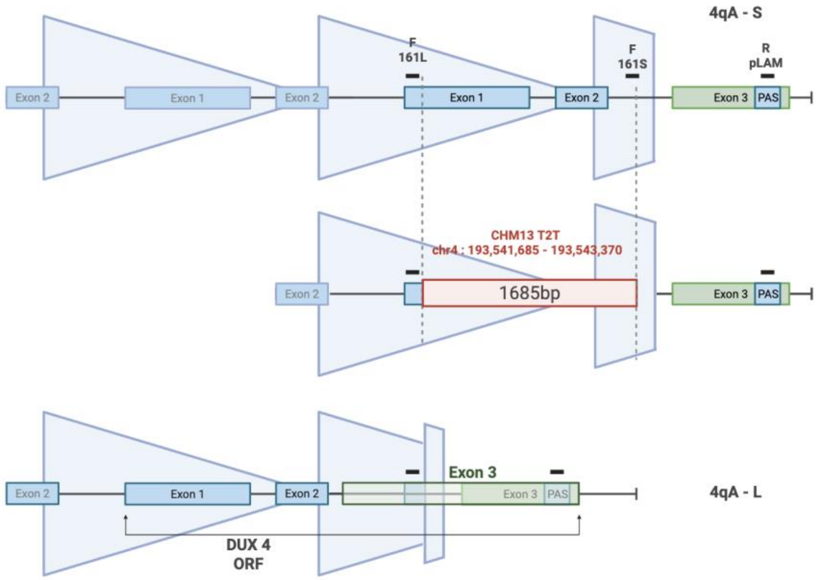

B

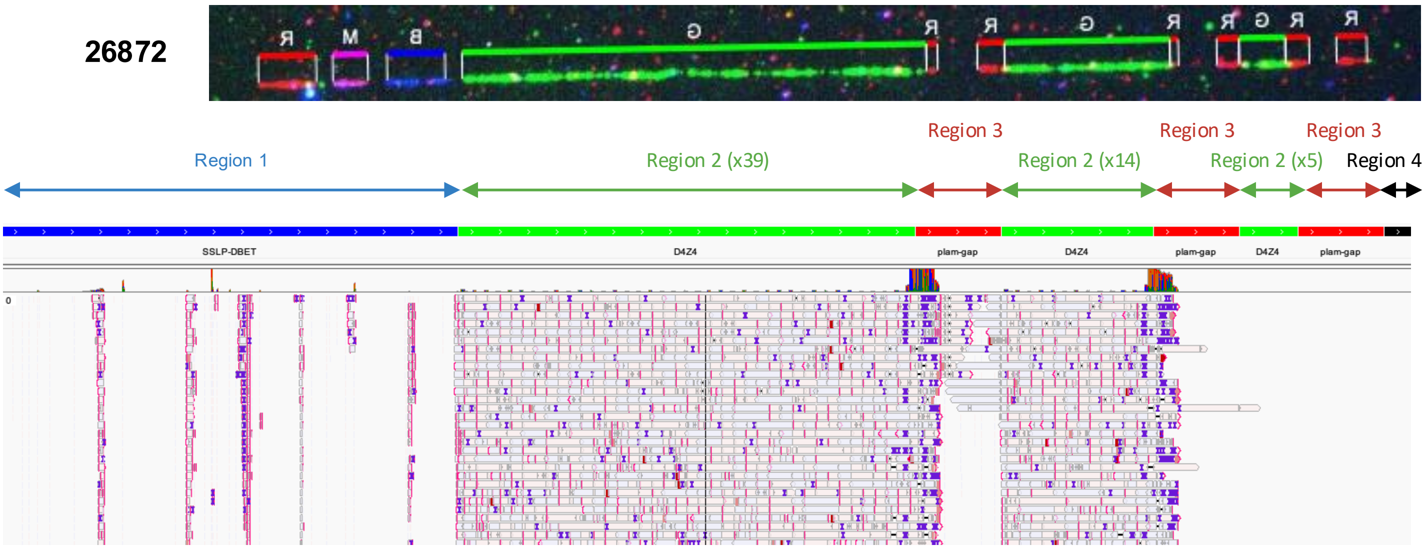

C

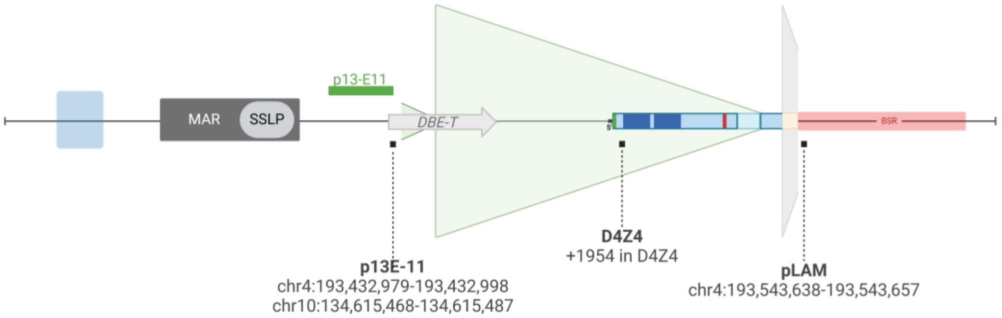

D

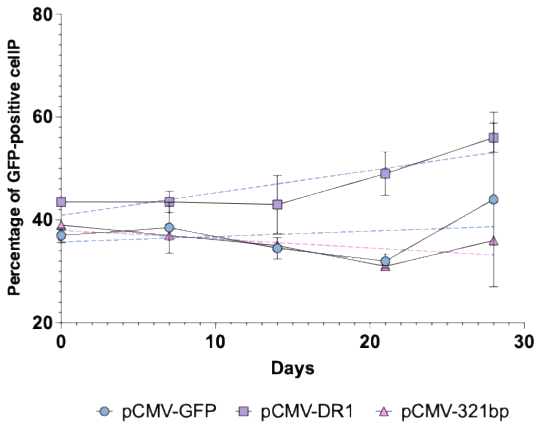

| | pCMV-GFP | pCMV-DR1 | pCMV-321bp |
|---|---|---|---|
| P value | 0.6747 | 0.0092 | 0.2452 |
| Deviation from zero? | Not Significant | Significant | Not Significant |
| Equation | Y = 0.1071*X + 35.70 | Y = 0.4357*X + 40.90 | Y = -0.1714*X + 38.00 |

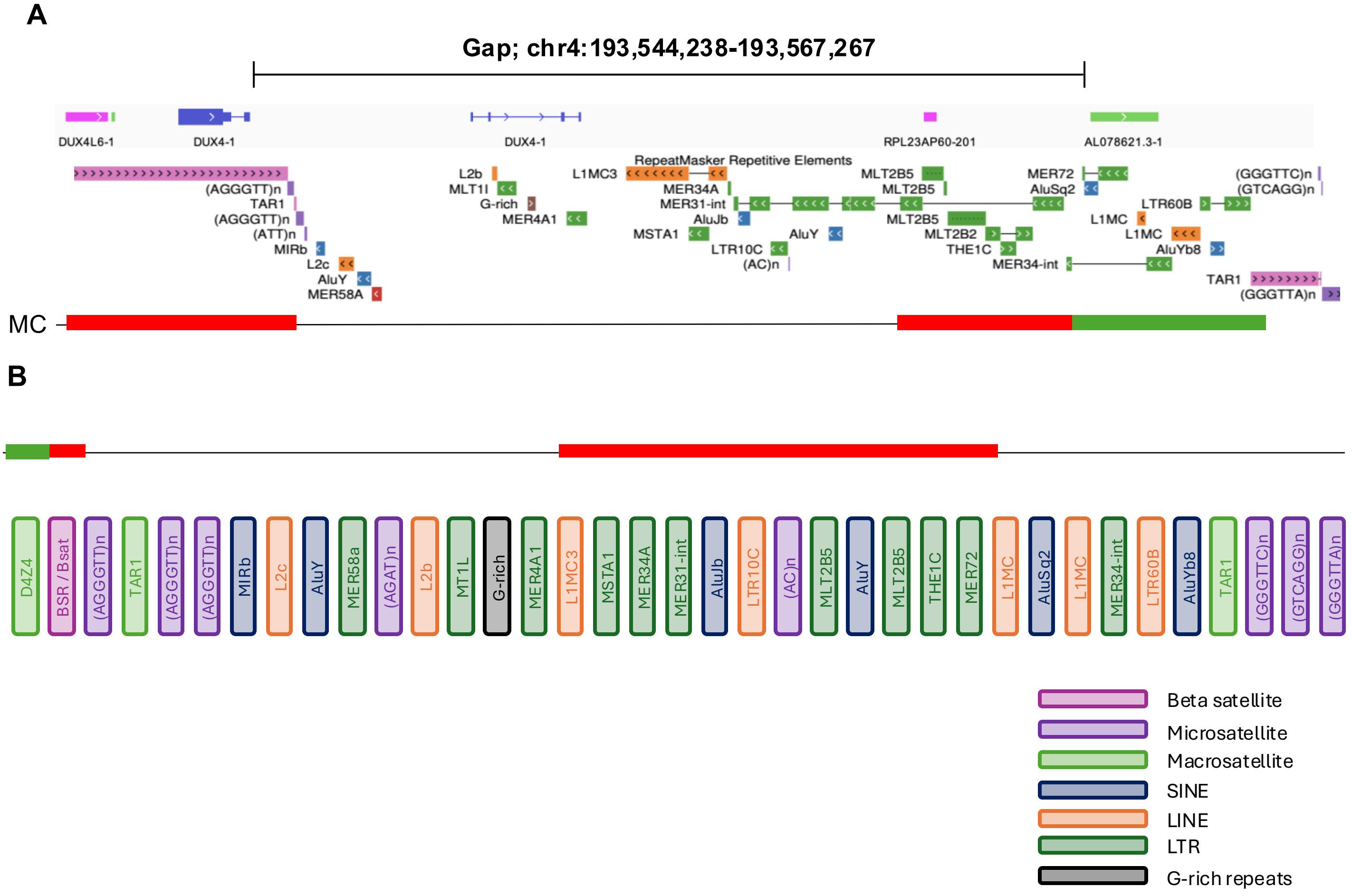

A
Gap; chr4:193,544,238-193,567,267
DUX4L6-1
DUX4-1
DUX4-1
RPL23AP60-201
AL078621.3-1
RepeatMasker Repetitive Elements
(AGGGTT)n
TAR1
(AGGGTT)n
(ATT)n
MIRb
L2c
AluY
MER58A
L2b
MLT1I
G-rich
MER4A1
L1MC3
MER34A
MER31-int
AluJb
MSTA1
LTR10C
(AC)n
AluY
MLT2B5
MLT2B5
MLT2B5
MLT2B2
THE1C
MER34-int
MER72
AluSq2
L1MC
L1MC
LTR60B
AluYb8
(GGGTTC)n
(GTCAGG)n
TAR1
(GGGTTA)n
MC
B
D4Z4
BSR / Bsat
(AGGGTT)n
TAR1
(AGGGTT)n
(AGGGTT)n
MIRb
L2c
AluY
MER58a
(AGAT)n
L2b
MT1L
G-rich
MER4A1
L1MC3
MSTA1
MER34A
MER31-int
AluJb
LTR10C
(AC)n
MLT2B5
AluY
MLT2B5
THE1C
MER72
L1MC
AluSq2
L1MC
MER34-int
LTR60B
AluYb8
TAR1
(GGGTTC)n
(GTCAGG)n
(GGGTTA)n
Beta satellite
Microsatellite
Macrosatellite
SINE
LINE
LTR
G-rich repeats

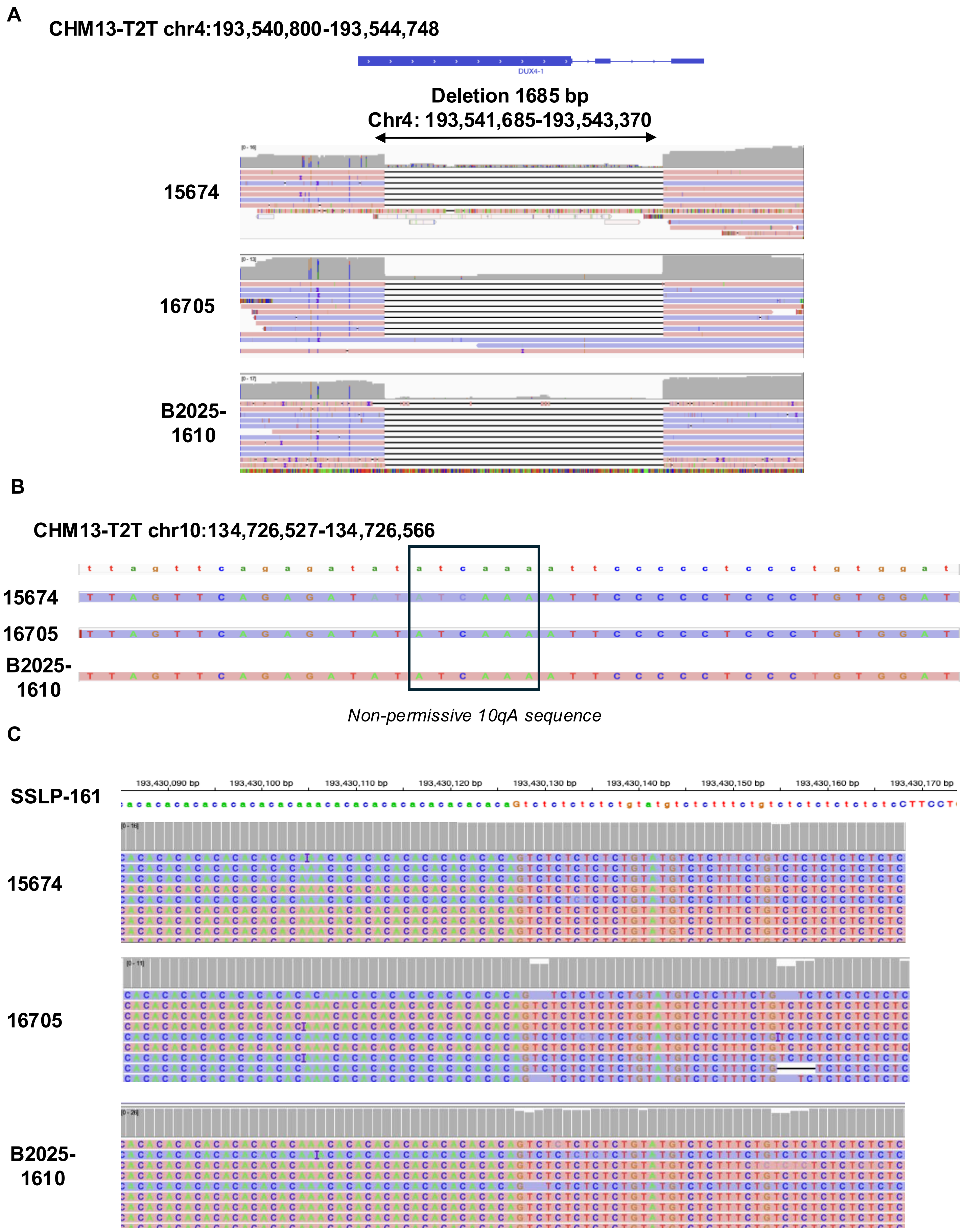

Tardy *et al* Supplementary figure 5

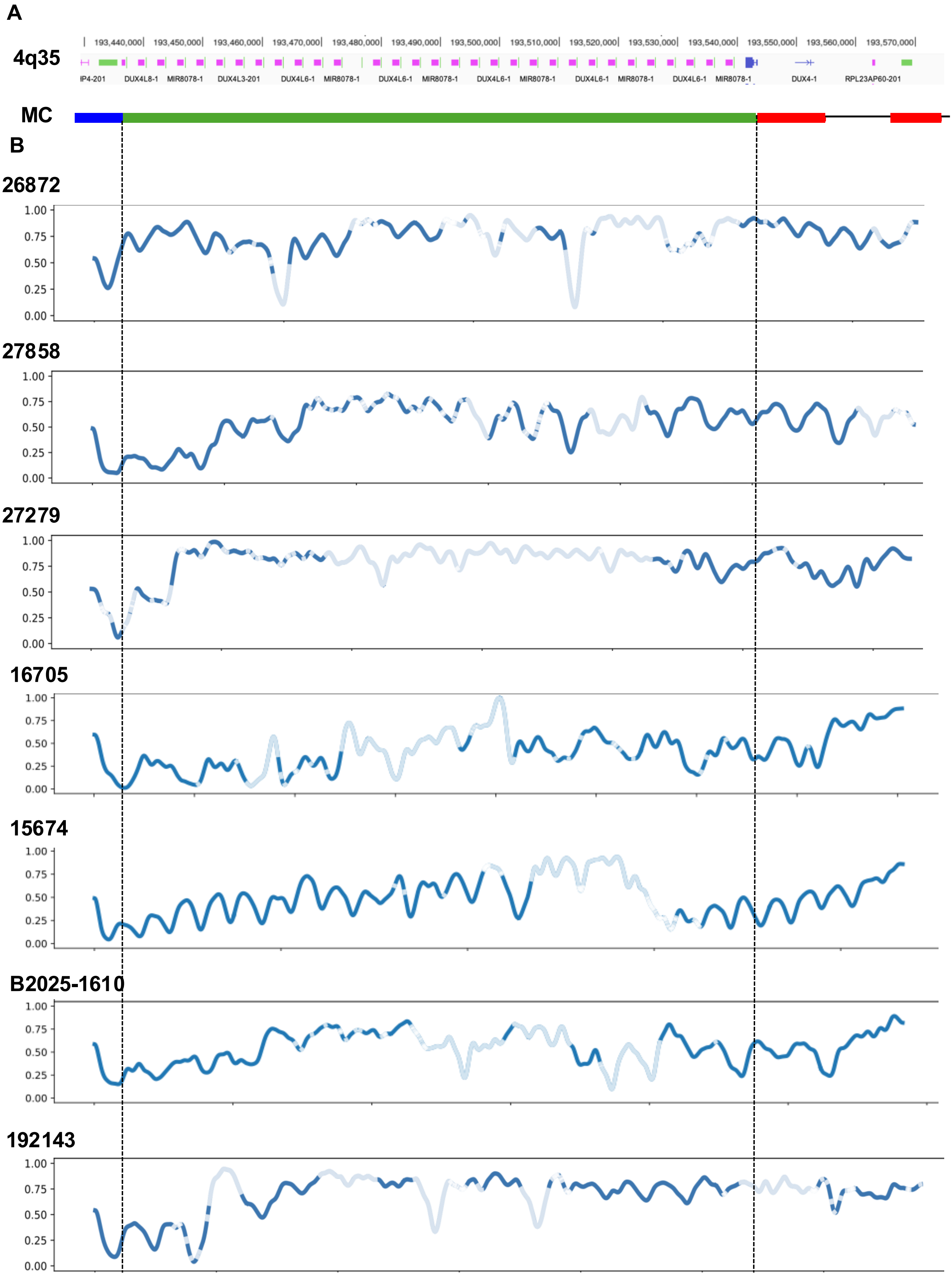

Tardy *et al*

Supplemental Figure 6

### 4q last D4Z4 - chr4:193,539,766-193,543,060

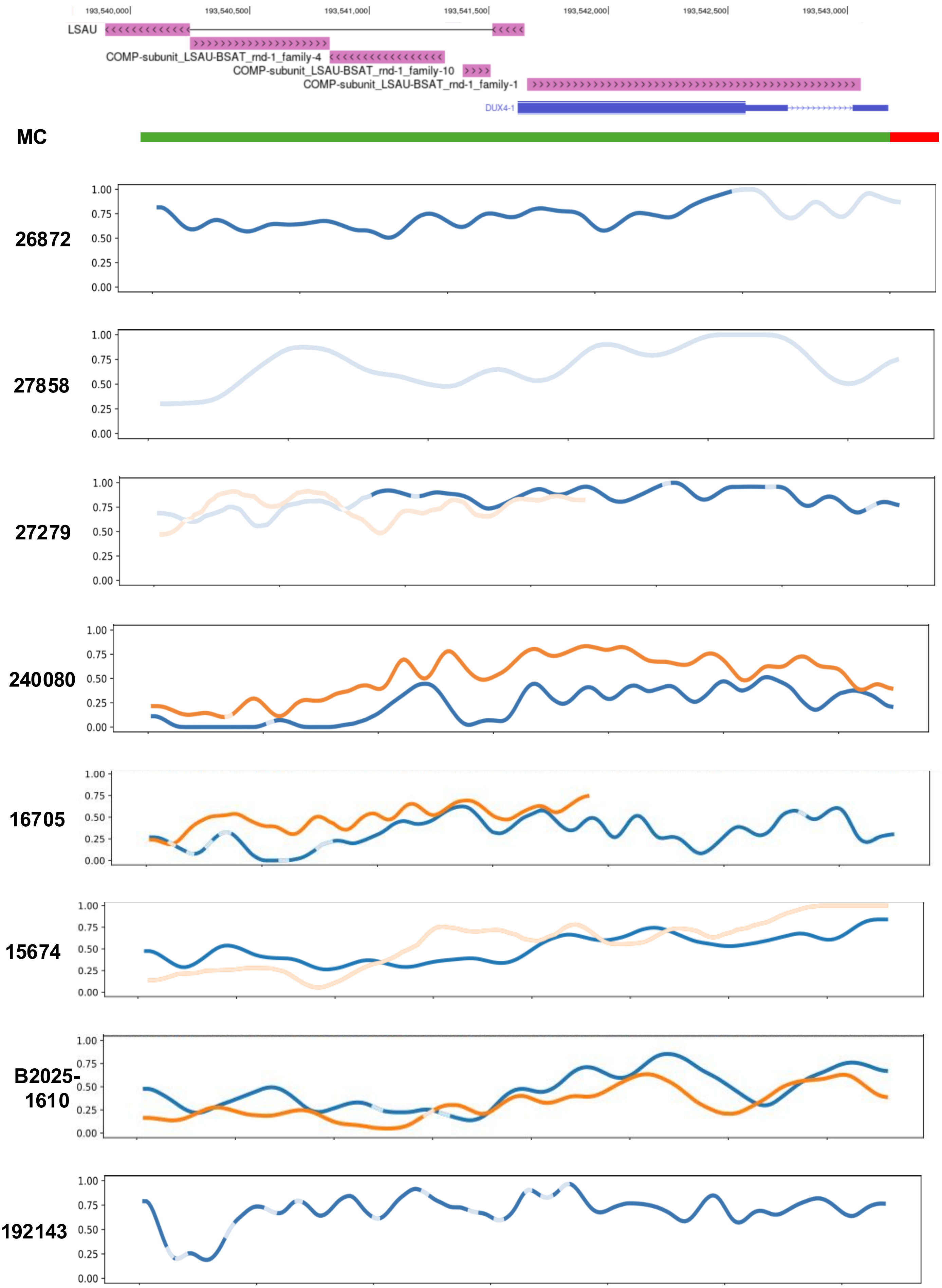

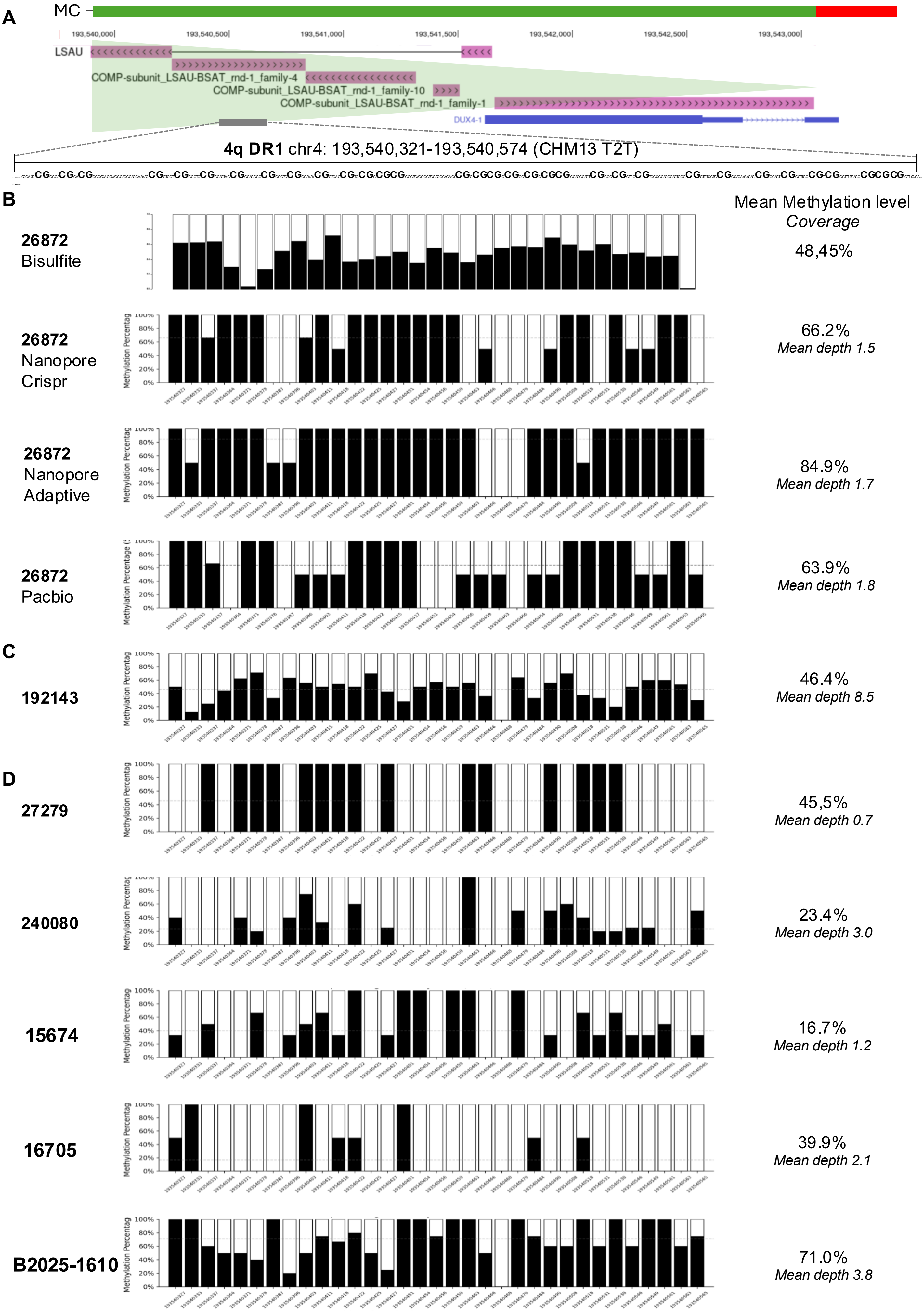

Tardy *et al*

**Supplemental Figure 8**

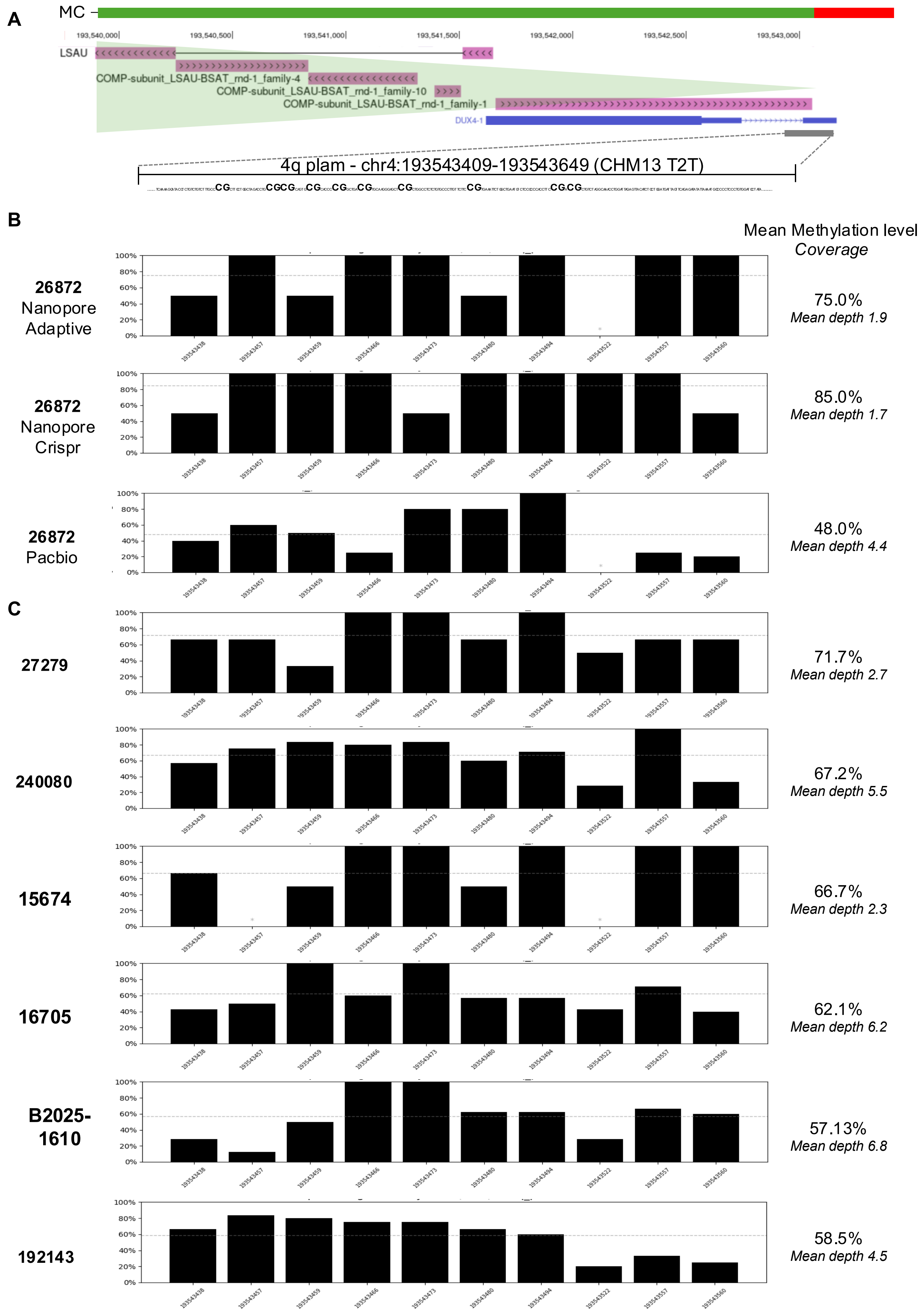

Tardy *et al*

**Supplemental Figure 9**
