## Supplemental information for "Comparison of long-read sequencing strategies for resolving complex genotypes at Facioscapulohumeral dystrophy-associated loci"

**Supplementary information**

**Materials and Methods**

### **Patients and samples.**

Individuals explored in this study were clinically assessed by neurologists with expertise in neuromuscular diseases who defined the presence or absence of clinical signs and evaluated the involvement of the groups of muscles typically affected in the disease (facial, shoulder and pelvic girdle, upper and lower limbs, and abdominal muscles) and level of impairment. Patients 27858; 240080, 15674, 16705 and B2025-1610 display clinical signs of FSHD. Patient 240080 carries a pathogenic variant in *SMCHD1* (NM_015295.3:c.1295T>C: p.(Phe432Ser)) located within the ATPase domain. Patients 15674, 16705 and B2025-1610 also carry a pathogenic variant in *SMCHD1* (NM_015295.3; Exon 28, c.3631C>T; p.Q1211*). Patient 26872 shows signs of muscular dystrophy and FSHD testing was part of a differential diagnosis procedure. For patient 27279 displays a mild muscular weakness and essential tremor and FSHD was part of a differential diagnosis for this patient (Table S2). Blood cells were embedded in agarose plugs for high-molecular-weight DNA extraction. For all patients, DNA was analyzed prior with MC (1). D4Z4 methylation level was assessed by Sodium Bisulfite Sequencing (BSS) (2) and patients were screened for the presence of a variant in *SMCHD1* (3-5). Informed consent was obtained from all patients for the genetic analyses (Table S2).

Samples were provided by the Center for biological Resources (Department of Medical Genetics, La Timone Children’s hospital) with the AC-2011-1312 (sample collection) and N°IE-2013-710 (ethical approval, Assistance Publique des hôpitaux de Marseille) accreditation numbers. Informed consent was obtained for all patients participating in the study or legal guardian for minor participants.

#### **DNA Extraction.**

For control 192143 and patient 27858 DNA was isolated and purified from agarose plugs used for MC, according to the New England Biolabs (NEB) #M0392 protocol (https://www.neb.com/en/protocols/0001/01/01/dna-purification-from-agarose-gels-using-beta-agarase-i-m0392). The length of the isolated DNA was determined by electrophoresis on a 0.4% agarose gel using the GeneRuler High Range DNA Ladder. DNA smears were always above 48kb. For all other samples, high molecular weight genomic DNA was purified from whole blood using Qiagen FlexiGene DNA Kit according to manufacturer’s instructions.

### **Targeted Nanopore Sequencing**

Multiple guide RNAs (gRNAs or crRNA) were used to target the p13E-11 proximal region (5’- CCT ATT AAA CGT CAC GGA CAG TTT TAG AGC TAT GCT-3’), the distal pLAM (5’- AAA TCT TCT ATA GGA TCC ACG TTT TAG AGC TA-3’), or D4Z4 (5’- CAC CAG AGA ACG GCT GGC CCG TTT TAG AGC TAT GCT -3’). For patient 26872 and control 192143, only the D4Z4 guide was initially used in order to specifically sequence the duplicated region and the gap (region that is not covered by MC probes) between duplicated arrays, for patient 27858, the pLAM, p13-E11 and D4Z4 guides were used (Table S3).

For the Cas9 targeting approach, sequencing was performed on a MinION R9.4.1 flow cells and ran on a MinION Mk1B after a ligation step with the SQK-CS9109 ONT Cas9 Sequencing Kit according to manufacturer’s instructions. Libraries were prepared by the Genomics and Bioinformatics facility (GBiM) at the Marseille Medical Genetics lab. Briefly, 1 μL of crRNA (either individually or pooled in equal volumes) was mixed with 1 μL of 100 μM tracrRNA (IDT #1072532) and 8 μL Duplex buffer, then heated at 95°C for 5 minutes. To assemble the Cas9 ribonucleoprotein complexes (RNPs), 23.7 μL of nuclease-free water, 3 μL of reaction buffer, 3 μL of the annealed crRNA, and 0.3 μL HiFi Cas9 Nuclease v.3 (IDT #1081060) were mixed in a 1.5 mL Eppendorf tube and incubated at room temperature for 30 minutes. For genomic DNA dephosphorylation, 3 μL of reaction buffer (RB), 24 μL of genomic DNA (1-5 μg depending on the sample), and 3 μL of phosphatase were incubated at 37°C for 10 minutes, then heated at 80°C for 2 minutes. Cleavage and dATP-tailing of the dephosphorylated DNA was performed by adding 10 μL of the Cas9 RNPs, 1 μL of 10 mM dATP, 1 μL of Taq polymerase, and incubating at 37°C for 15 minutes, followed by a 5-minute incubation at 72°C. Sequencing adapters were ligated to the genomic DNA by mixing 20 μL ligation buffer (LNB), 3 μL nuclease-free water, 10 μL T4 DNA ligase (LIG), and 5 μL adapter mix (AMX) in a new clean Eppendorf tube. This mixture was added to the sample in two steps of 20 μL and 18 μL, then incubated at room temperature for 10 minutes. The Cas9 library was purified using AMPure XP beads (Beckman Coulter #A63881) by adding 80 μL of SPRI dilution buffer to the 80 μL library, binding it to 48 μL of beads at room temperature for 10 minutes, washing the beads twice with 250 μL of short fragment buffer, briefly drying, and resuspending them in 13 μL of elution buffer (EB). This suspension was incubated at room temperature for 30 minutes before being transferred to a new clean Eppendorf tube. The MinION Flow Cells were primed with 800 μL of flush buffer (FB) with previously added flush tether (FLT). For loading, 12 μL of the Cas9 libraries (200fmol) were mixed with 37.5 μL of sequencing buffer (SQB) and 25.5 μL of loading beads (LB), then loaded onto the MinION Flow Cell. Data collection was managed using MinKNOW 23.04.6 software on a PC desktop computer via a USB 3.0 port.

### **Nanopore Adaptive sampling sequencing**

Sequencing was performed on a MinION FLO-MIN114 flow cell mounted on a GridION Mk1 device using the SQK-LSK114 ligation sequencing kit (Oxford Nanopore Technologies), following the manufacturer’s instructions. Two libraries were prepared from 7,1 µg of high-molecular-weight genomic DNA. Briefly, DNA was subjected to end-repair and dA-tailing by adding 7 µL of NEBNext FFPE DNA Repair Buffer, 3.5 µL of FFPE DNA Repair Mix, 3.5 µL of NEBNext Ultra II End Repair / dA-Tailing Buffer, and 3.5 µL of End Prep Enzyme Mix (New England Biolabs), and incubating the mix at 20°C for 5 minutes followed by 65°C for 5 minutes. The DNA was then cleaned up using a 1x volume of AMPure XP beads, washed twice with 80% ethanol, and eluted in 60 µL of nuclease-free water. Adapter ligation was performed by adding 25 µL of ligation buffer (LNB), 10 µL of NEBNext Quick T4 DNA Ligase, and 5 µL of Adapter Mix II (AMX) to the repaired DNA, followed by a 10-minute incubation at room temperature. After ligation, the sample was purified using 0.4x AMPure XP beads, washed with Short Fragment Buffer (SFB), and eluted in 15 µL of elution buffer (EB). Prior to sequencing, 12 µL (200fmol) of the library were mixed with 37.5 µL of sequencing buffer (SQB) and 25.5 µL of loading beads (LB) and loaded onto the flow cell. Adaptive sampling was performed using a custom reference HG002 diploid genome reference (hg002v1.0.1.fasta) and a custom BED file targeting the 4qA and 4qB subtelomeric regions (Supplementary Code). Data collection was managed using MinKNOW v24.06.14 with adaptive sampling enabled.

### **Nanopore Whole Genome sequencing**

For whole genome sequencing, 3,7 µg of high-molecular-weight genomic DNA were used to prepare two libraries, which were sequentially loaded with washes performed between each load on a FLO-PRO114M PromethION flow cell using the SQK-LSK114 ligation sequencing kit (Oxford Nanopore Technologies), according to the manufacturer’s instructions. Sequencing was performed on a PromethION 2 Integrated. Briefly, DNA was subjected to end-repair and dA-tailing in a total volume of 69.5 µL by adding NEBNext FFPE DNA Repair Buffer and Mix, NEBNext Ultra II End Repair / dA-Tailing Buffer, and End Prep Enzyme Mix, with incubation at 20°C for 5 minutes followed by 65°C for 5 minutes. The reaction was purified using a 1x AMPure XP bead clean-up, washed twice with 80% ethanol, and eluted in 60 µL nuclease-free water. Adapter ligation was carried out by adding 25 µL of ligation buffer (LNB), 10 µL of NEBNext Quick T4 DNA Ligase, and 5 µL of Adapter Mix II (AMX), with a 10-minute incubation at room temperature. Following adapter ligation, DNA was purified with 0.4x AMPure XP beads, washed with SFB, and eluted in 15 µL of elution buffer. For each load, 12 µL (200fmol) of the library were mixed with 37.5 µL of sequencing buffer and 25.5 µL of loading beads and then loaded onto the flow cell. Washes were performed between each loading step according to ONT recommendations. Sequencing runs were monitored and controlled using MinKNOW v25.03.7.

**Bionano Optical Genome Mapping**

Optical genome mapping was performed using the Bionano Saphyr system with the Direct Label and Stain (DLS) method, following the manufacturer’s protocols. Briefly, ultra–high molecular weight genomic DNA was isolated and labeled using the DLE-1 enzyme, which introduces fluorescent tags at specific sequence motifs across the genome. The labeled DNA molecules were loaded onto a Bionano Saphyr Chip and imaged on a Saphyr instrument. DNA molecules were linearized in nanochannels and fluorescently imaged to generate high-resolution optical maps. The data were analyzed using Bionano Solve software and the EnFocus™ FSHD Analysis pipeline, which is specifically designed to detect D4Z4 repeat contractions and distinguish 4qA from 4qB alleles, as well as assess 10q homologous regions.

### **Pacbio circular Consensus sequencing**

6 µg high-molecular-weight genomic DNA was fragmented to ~30 kb. Library preparation was performed using the SMRTbell® Express Template Prep Kit 2.0 (Pacific Biosciences). Sequencing was performed on a PacBio Revio system using SMRT Cell. Basecalling of sequenced bases and methylated bases was managed using SMRT Link v13.1.0.221970. High-fidelity (HiFi) reads were generated using the Circular Consensus Sequencing (CCS) mode with default parameters. Reads were aligned to the T2T-CHM13 v2.0 reference genome using pbmm2 (v1.9.0) with the --preset CCS option. Sequencing was performed on the Gentyane core facility, UMR INRAE/UCA 1095 GDEC, Clermont Ferrand, France.

### **Basecalling of modified bases and alignment.**

Basecalling and alignment were performed using epi2me wf-human-variation workflow (<https://github.com/epi2me-labs/wf-human-variation>). The raw sequencing data in FAST5 format were converted to POD5 format using Dorado. Basecalling of modified bases (5mCG), was performed on the converted POD5 files using the Dorado (https://github.com/nanoporetech/dorado) basecaller. The basecalled bases were redirected to a BAM file and aligned to the reference genome (CHM13-T2T v2.0 or HG002) or custom reference using the Dorado aligner. The aligned BAM file was sorted and indexed using Samtools.

### **DNA Methylation Analysis**

The .bam files were processed with Modkit (https://github.com/nanoporetech/modkit) to extract base modification data. The profile of methylated CpG along the D4Z4 unit was generated using Methylartist version 1.5 (<https://github.com/adamewing/methylartist>). Methylartist processes BAM files containing modification tags (MM/ML), enabling visualization of methylation patterns at a single-molecule resolution, as well as aggregated methylation frequencies across the targeted genomic locus, focusing on CpG motifs.The specific methylation histograms for the DR1 and pLAM sequence were produced using a custom Python script available in the supplementary code.

**Quantitative RT-PCR**

Reverse transcription of 1 µg of total RNA was performed using the Superscript IV First-Strand cDNA Synthesis kit (Life Technologies) with a mix of oligo dT and random hexamers. Primers were described in (6). Quantitative PCR comply with the MIQE guidelines.

**Neuromuscular differentiation**

The hIPSc were differentiated into functional muscles according to the protocol described in (7). Cells were collected 30 days post differentiation.

**RNA extraction, quality control and library preparation**

Total RNA was extracted using the RNAeasy kit (Qiagen) following the manufacturer’s instructions.

**Legends to the supplementary figures**

**Figure S1. Comprehensive analysis of distal 4q and 10q subtelomeres using the CMH13 T2T genome assembly.**

**A.** We positioned specific 4q- and 10q-associated genomic regions (p13E11, proximal SSLP, D4Z4, pLAM) relative to the T2T reference genome using chm13v2.0.fa and the probes used for MC relative to this genome assembly. The CHM13-T2T assembly has the 4A161S variant and contains an ATTAAA polyadenylation signal on chromosome 4 (chr4:193,543,619-193,543,624). The homologous region between 10q and 4q chromosomes begins distal to an inverted partial D4Z4 unit, located ~46 kb upstream the D4Z4 array on 4q, at locations chr4:193,391,133 and chr10:134,573,992. Different sequence length polymorphism (SSLP) with four major haplotypes: 161, 163, 166, and 168 located in a Matrix Attachment Domain (MAR) were described in the proximal part of 4q and 10q loci. **B.** The region corresponding to the p13E11 probe (D4F104S1 marker, 820 bp long) is located upstream for the first D4Z4 unit and hybridizes at the 5’ end of the first truncated D4Z4, which contains the end of a D4Z4 unit, a 3’ end fragment of the COMP-subunit_LSAU_BSAT_rnd-1_family-1, 887bp-long, with a deletion of 564bp in its 5’ part, starting at chr4:193,433,358. This assembly was annotated for repetitive sequences of the Dfam 3.3 database that were aligned with RMBlast. The resulting data are available on the UCSC Genome Browser under the RepeatMasker track**. C.** Schematic representation of a D4Z4 unit. Each D4Z4 begins with a “GATC” sequence within a LSau element. LSau repeats are long (400-500 bp) Sau3A DNA repeats with higher GC content than typical β-satellite repeats (68% vs 54%). The *Kpn*I restriction site is located +23nt within the LSau. RMBlast aligns Lsau in CHM13-T2T in antisense and interrupted by composite Lsau/βsatellite sequences. This results in a first LSau fragment 367nt long, and a second 138nt long, in the middle of D4Z4. **D.** In the distal part of the locus, the pLAM sequence starts with an 501nt LSau sequence, 99% similar to the D4Z4 repeat until around nucleotide 348, where a unique sequence specific to pLAM begins. The distinction between the 4qA, 10qA and 4qB sequences lies in the presence of a 6.4-kb β-satellite of 68 bp Sau3A tandem repeats (CHM13-T2T; chr4:193,543,567-193,550,04). DUX4 exon 3 starts at nt392 in the Lsau and continues 84bp into the β-satellite.

**Figure S2. Low Mapping Quality to Custom Reference in Patient 26872.**

**A.** Raw MC image for control 192143. From left to right: the proximal red probe is specific to chromosome 4. The pink probe maps the proximal region that is shared between chromosomes 4 and 10. The blue probe maps to the p13E11 region. The green probe corresponds to D4Z4. Downstream of the green probe, the blue probe corresponds to the qB haplotype. The D4Z4 array contains 14 repeated D4Z4 units (RUs). The most distal red probe on the right corresponds to the distal part of the 4q35 subtelomeric region**. B.** Off target sequencing of D4Z4-like elements. Plots showing the sequencing coverage of ONT CRISPR reads distributed across the entire genome, including off-target regions.

**Figure S3**. **Position of CRISPR guides for ONT sequencing.**

**A**. Schematic representation of D4Z4 and distal pLAM regions. The position of the polyadenylation site (PAS) is indicated together with the position of PCR primers used distinguish short from long alleles (8): 4AS-F (286bp) 5′-CCC GCC CGG GCC CCT GCA-3′ (CHM13-T2T chr4:193,543,358-193,543,375) ; 4AL-F (330bp) 5′-CGA GGA CGG CGA CGG AGA C -3′ (CHM13-T2T chr4:193,541,630-193,541,647) reverse primer PAS-R 5′-GAT CCA CAG GGA GGG GGC ATT TTA-3′ (CHM13-T2T chr4:193,543,621-193,543,644) (upper panel). 4qL distal haplotype differs from 4qA-S in the form of a 1685bp loss of part of the last D4Z4 and pLAM which contains the 161S primer (middle panel) that is deleted when reads are aligned relative to the CHM13-T2T sequence of reference which is 4qA-S (positions chr4:193, 541, 685-193,543,370). This explains that only the 161L PCR sequence is produced in 4qA-L patients (bottom panel). **B.** Alignment of ONT reads for patient 26872, who carries a 4qA *cis*-triplicated allele (39 + 14 + 5 RUs) as determined by MC. From top to bottom, MC image, the reconstructed reference corresponding to the patient's 4qA allele, and the alignment of sequencing reads visualized in IGV. **C.** Position of the different CRISPR guides relative to the CHM13-T2T genome assembly and mapping to p13E11, D4Z4 and the distal pLAM sequence (Table S3) (dashed lines).

**Figure S4.** **Simplified representation of the repeated sequences spanning the 4q35 gap region.**
**A.** We analyzed the sequence composition of the region comprised between *cis*-duplicated arrays and that is not covered by MC probes (Gap; chr4:193,544,238-193,567,267 relative to CHM13-T2T). Upper panel: ordered succession of repeated elements grouped by category and colored accordingly: Beta satellite (*BSR/Beta*, pink), microsatellites (*AGGGTT)n, (ATT)n, (AGAT)n, (AC)n, (GGGTTC)n, (GTCAGG)n, (GGGTTA)n*, purple), macrosatellites (*TAR1, D4Z4*, green), SINEs (*AluY, AluJb, AluSq2, AluYb8, MIRb*, blue), LINEs (*L2b, L2c, L1MC, L1MC3*, orange), LTRs (*MER58A, MLT1L, MER4A1, MSTA1, MER34A, MER31-int, LTR10C, MLT2B5, THE1C, MER72, LTR60B, MER34-int*, dark green), and G-rich repeats (*G-rich*, grey). **B.** closeup view of the molecular combing pattern of the cis-duplicated region.

**Figure S5. Nanopore sequencing complements Bionano Optical Genome Mapping in case of complex genotypes**

**A.** Close-up view of the *DUX4* sequence. For all three patients (15674, 16705, B2025-1610), the 4qA-L haplotype that corresponds to the presence of a longer final intron, is visible as a 1685 bp deletion when reads are compared to the T2T reference genome. **B.** Sequence analysis of the last D4Z4 repeat on chromosome 10 showing the presence of a non-permissive pLAM ATCAAA sequence (chr10:134,726,527-134,726,566) instead of the 4qA ATTAAA sequence for all three patients. **C**. IGV view of the SSLP polymorphism at the 4qA locus shows a homozygous SSLP-161 allele (CA10 AA CA10 GT CT5 GT AT GT CT2 TT CT GT CT6) for all three patients.

**Figure S6. Methylation analysis of the D4Z4 region obtained by ONT sequencing.**

**A.** IGV view of the 4q35 region relative to the CMH13-T2T genome assembly (upper panel; chr4:193,429,551–193,574,945; CHM13-T2T). MC bar code from the p13E11 probe (blue) to the most distal telomeric probe (red). The green probe corresponds to D4Z4 **B.** Methylation profile across the entire 4q35 locus (chr4:193,429,551–193,574,945; CHM13-T2T), obtained using methylartist v1.5. From top to bottom: RefSeq track from UCSC, position of the MC probes (from left to right, p13E11 (Blue), D4Z4 (green), A-type haplotype (red) and distal telomeric probe (red)), methylartist methylation plot with the methylation profile visualized in blue (*x-axis*: chromosomal position, *y axis*: mean methylation percentage) for patients 26872, 27858, 27279, 16705, 15674, B2025-1610 and control (192143). Light blue corresponds to regions with a low depth of coverage. The region marked by dashed lines corresponds to the D4Z4 array.

**Figure S7. Methylation profile of the last D4Z4 unit at 4q obtained by ONT sequencing.**

**A.** IGV view of the last D4Z4 unit and flanking A-type haplotype for the 4q35 region relative to the CMH13-T2T genome assembly (upper panel; chr4:193,539,766-193,543,060). The position and identity of the different repetitive elements contained within this region and identified by Repeat masker are indicated, together with the *DUX4* coding sequence (blue). Lower panel, MC bar code for D4Z4 (green) and A-type haplotype (red). **B**. methylartist methylation plot. Depending on the depth of coverage, different haplotypes might be identified and visualized by in blue or orange. Regions of low depth are represented in light blue or light orange. *x-axis*: chromosomal position, *y axis*: mean methylation percentage for patients 26872, 27858, 27279, 240080, 16705, 15674, B2025-1610 and control (192143).

**Figure S8. Comparisons for DNA methylation profiling at DR1 across different methods.**

**A.** Upper panel, MC bar code for D4Z4 (green) and A-type haplotype (red). IGV view of the last D4Z4 unit and flanking A-type haplotype for the 4q35 region relative to the CMH13-T2T genome assembly (upper panel; chr4:193,539,766-193,543,060). The position and identity of the different repetitive elements contained within this region and identified by Repeat masker are indicated, together with the *DUX4* coding sequence (blue) and the CpG dinucleotides contained within this region. **B-D.** For each histogram bar, black corresponds to the percentage of methylated CG and cumulated white bar, to the percentage of unmethylated CG. **B.** Sodium bisulfite sequencing at DR1 (31 CpGs, 48.45%) for patient 26872 compared to the methylation profile at DR1 for the most distal D4Z4 unit obtained by ONT adaptive sampling (33 CpGs; 84.9%), ONT targeted sequencing (33 CpGs; 66.2%) and Pacbio sequencing (33 CpGs; 63.9%). **C.** ONT Methylation profile at DR1 for the most distal D4Z4 unit for control 192143 (33 CpG; 46.4%). **D.** ONT Methylation profile at DR1 for the most distal D4Z4 unit for patient 27279 (33 CpG; 45.5%), patient 240080 (33 CpG; 23.4%)**,** patient 16705 (33 CpG; 39.9%)**,** patient 15674 (33 CpG; 16.7%)**,** patient B2025-1610 (33 CpG; 71%). For all conditions, individual CpGs are shown on the x-axis and modification percentage of methylated sequences on the y-axis. The global methylation level was calculated as the ratio of the number of methylated CGs relative to the total number of CG. The mean coverage depth is indicated on the right for each sample.

**Figure S9. DNA methylation profiling of the pLAM region in patients with complex rearrangements.**

**A.** Upper panel, MC bar code for D4Z4 (green) and A-type haplotype (red). IGV view of the pLAM region at 4q35 relative to the CMH13-T2T genome assembly (chr4: chr4:193,543,409-193,543,649). The position and identity of the different repetitive elements contained within this region and identified by Repeat masker are indicated, together with the ATTAAA polyadenylation site (PAS) and the CpG dinucleotides contained within this region. **B.** For each histogram bar, black corresponds to the percentage of methylated CG and cumulated white bar, to the percentage of unmethylated CG. Methylation profile of the pLAM region (Chr4: 193,543,409-193,543,649) in patient 26872 obtained by ONT adaptive sequencing (10 CpGs; 75.0%), nCATS (10 CpGs; 85.0%), and PacBio (10 CpGs; 48%). **C.** Methylation profile of the pLAM region obtained by ONT adaptive sampling or WGS for control 192143 (10 CpGs; 58.50%), patients 27279 (10 CpGs; 71.7%), 240080 (10 CpGs; 67.2%), 16705 (10GpGs, 62.1%), 15674 (10CpGs; 66.7%), and B2025-1610 (10CpG; 57.13%). Patient 27858 is not presented as the number of reads was not sufficient for DNA methylation analysis. For all conditions, individual CpGs are shown on the *x-axis* and the percentage of methylated sequences on the *y-axis*. The global methylation level was calculated as the ratio of the number of methylated CGs relative to the total number of CG. The mean coverage depth is indicated on the right for each sample.
